# AWaRe antibiotic prescribing for common acute infections in low-middle income countries – a patient level analysis using IQVIA prescriber surveys from Pakistan, Egypt and Indonesia

**DOI:** 10.1101/2025.11.03.25339383

**Authors:** Nam Nguyen, Sherry Mangla, Peter Stephens, Aislinn Cook, Mike Thorn, Zikria Saleem, Siswanto Agus Wilopo, Elizabeth Tayler, Mike Sharland, Koen B. Pouwels

## Abstract

**Background:** There is limited large-scale and high-quality data on antibiotic prescribing in low- and middle-income countries (LMICs), particularly in the private sector. Here we use healthcare surveys to assess antibiotic prescribing levels and the factors influencing prescribing decisions for common infections in primary care and outpatient settings, predominantly within the private sector, in Pakistan, Egypt, and Indonesia.

**Methods:** We analysed surveys completed by prescribers in Pakistan, Egypt, and Indonesia, collected in primary care and outpatient settings, predominantly within the private sector, by IQVIA between 2017 and 2021, namely IQVIA’s proprietary Medical Data Index (Medical Index of Pakistan (MIP), Egypt Medical Data Index (EMDI) and Indonesia Medical Data Index (IMDI)). IQVIA market research information reflects estimates of real world activity and should be treated accordingly. We evaluated antibiotic prescribing categorized by WHO AWaRe and Essential Medicines List (EML) classifications for common infections. We used mixed-effects regression analyses to identify factors influencing prescribing decisions.

**Results:** Among the 384,975 infection-related health consultation records analysed, antibiotics were prescribed overall in 82.0% of consultations in Pakistan, 81.2% in Egypt, and 69.1% in Indonesia. Watch antibiotics accounted for 70.2% of all antibiotic prescriptions in Pakistan, 52.9% in Egypt, and 53.6% in Indonesia. Non-WHO EML antibiotics accounted for 26.8% of prescriptions in Pakistan, 39.9% in Egypt, and 33.0% in Indonesia. Consultations for patients presenting with lower respiratory tract infections, urinary tract infections, multiple infections, or differentiated fever had higher odds of receiving any or a Watch antibiotic. Consultations by respiratory-related specialists in Pakistan and Egypt and by most specialities in Indonesia were more likely to receive Watch antibiotics.

**Interpretation:** Similar patterns of very high levels of total and Watch antibiotic prescribing for most common acute infections - including those that generally do not require any antibiotics – were identified among prescribers in primary care and outpatient settings within the private sector in Pakistan, Egypt, and Indonesia.

**What is already known on this topic:**

- Pakistan, Egypt, and Indonesia are densely populated lower-middle income countries (LMICs), with a significant burden of antimicrobial resistance (AMR). However, evidence on antibiotic prescribing patterns and their drivers in primary care, particularly in the private sector, is limited

**What this study adds:**

- Antibiotic prescribing for common infections in primary care in Pakistan, Egypt, and Indonesia was very high, with antibiotics given in 70–80% of patient consultations.
- Despite WHO AWaRe book guidance recommending Access antibiotics or no antibiotics for most of these conditions, Watch antibiotics were frequently prescribed.
- High antibiotic prescribing appeared to be the norm, with low variability across prescribers, infections, and specialisations;

**How this study might affect research, practice or policy:**

- Urgent interventions are needed to address antibiotic overprescribing, which appears to have become routine in primary care and outpatient private healthcare in LMICs.

## INTRODUCTION

Antibiotics play a major role in reducing the burden of bacterial infections. However, their success is threatened by increasing antimicrobial resistance (AMR).^2^ Overuse of antimicrobials is known to be a major driver of AMR in low- and middle-income countries (LMICs).^3^ Yet, understanding the extent of antibiotic overuse in LMICs is often limited by lack of data on antibiotic prescribing.^4^ A paucity of data is particularly apparent in primary care and private settings, which serve as the formal first point of care for most patients with minor illnesses.^5^ These data gaps lead to challenges in conducting research, and potentially poor implementation of interventions to optimise antibiotic use.^6^

Pakistan, Egypt and Indonesia are three densely populated LMICs in the Eastern Mediterranean and South-East Asia regions with a significant burden of AMR. Previous studies based on aggregated data or small surveys suggest that antibiotic consumption in these countries is high and that antibiotics are being prescribed for conditions that generally do not require antibiotics. Antibiotic consumption in Pakistan increased by 65% between 2000 and 2015, positioning this country as the fourth highest consumer of antibiotics among LMICs in 2015, when adjusted for population size. Indonesia experienced an estimated 2.5-fold increase in antibiotic consumption over the same period, largely driven by an increase in the use of broad-spectrum antibiotics.^7–9^ Although comparable statistics are not available for Egypt, a small-scale survey showed that 64% of prescribers prescribed antibiotics to treat colds, and 21% acknowledged having prescribed antibiotics unnecessarily.^10^

While previous studies indicated that antibiotic overuse is common, a more comprehensive analyses on levels and patient- and prescriber-level drivers of antibiotic prescribing practices is needed to generate the evidence base required for the design and implementation of effective antibiotic stewardship interventions. We therefore analysed data from large-scale prospectively collected surveys submitted by prescribers in 18 geographic regions of Pakistan, Egypt and Indonesia.

## METHODS

### Data sources and procedures

We used three databases within the IQVIA Medical Data Index (Medical Index of Pakistan (MIP), Egypt Medical Data Index (EMDI), Indonesia Medical Data Index (IMDI)) for the period 2017-2021, which contain data on patient encounters prospectively collected by a representative sample of general practitioner and specialist prescribers, predominantly from the private sector, in Pakistan, Egypt and Indonesia. Data are collected in a standardised manner ensuring that data can be compared between countries. ^11^

In each country, IQVIA established a sampling frame to represent the national distribution of prescribers, recruiting doctors from various regions and specialties. Data collection, which encompassed patient encounters from all practice settings except in-patient care, covered any reason for seeking medical care. Trained recruiters approached prescribers. Eligible prescribers who agreed to participate were then instructed to record details of every patient encounter during a pre-determined week each quarter or semester throughout the year. Prescribers reported data from all patient encounters, regardless of whether a prescription was issued. The data requested for entry included patient demographic information, diagnosis, and treatment details (if any) in free text. IQVIA then coded diagnoses according to ICD-10 and classified prescribed medications using the European Pharmaceutical Research Association (EphMRA) Anatomical Therapeutic Classification (ATC) system.^12,13^ For all antibiotic products, we converted the EphMRA ATC codes to WHO ATC codes, and added information on WHO AWaRe and Essential Medicines List (EML) classifications.^14,15^ Prescriber and practice characteristics, including prescriber’s sex, specialty, and practice setting (hospital or clinic or others), were also collected from prescribers. Only aggregated data were transferred from IQVIA.

### Participants

We obtained health records for outpatient consultations related to common acute infections in Pakistan, Egypt, and Indonesia between 2017 and 2021 (Appendix 1). All records were related to outpatient consultations in the private sector except in Indonesia, where 6.6% were from outpatient departments of government hospitals (Table 1). These records were submitted by doctors across 8 regions in Pakistan, 4 regions in Egypt, and 6 regions in Indonesia. Visits were included if they had an ICD-10 code corresponding to an acute common infection diagnosis, falling within one of the following groups: respiratory tract infections (RTIs), gastrointestinal infections (GIs), urinary tract infections (UTIs), ear infections, skin infections, or fever of unknown origin. The list of acute common infections was initially developed through a literature review of published research on similar topics.^16^ To ensure that no relevant acute infections were missed, we extracted diagnoses and ICD-10 codes from all visits associated with antibiotic prescriptions from the datasets. We added any conditions not initially included in the original list but deemed relevant upon review into the final list (Appendix 2).

**Table 1.** Demographic and health-related characteristics of patients and prescribers by country.

|  | **Pakistan**<br>**(n=204 822)** | **Egypt**<br>**(n= 87 286)** | **Indonesia**<br>**(n=92 807)** |
| --- | --- | --- | --- |
| **Age, median [IQR]** | 25 [10-42] | 26 [07-43] | 27 [10-42] |
| **Age group** |  |  |  |
| ≤ 5 | 31 153 (15.2) | 19 738 (22.6) | 14 417 (15.5) |
| 6 – 10 | 15 152 (7.4) | 7 607 (8.7) | 6 499 (7) |
| 10 – 17 | 21 054 (10.3) | 6 705 (7.7) | 7 169 (7.7) |
| 18 – 24 | 28 728 (14.0) | 8 341 (9.6) | 11 014 (11.9) |
| 25 – 39 | 59 254 (28.4) | 18 994 (21.8) | 24 833 (26.8) |
| 40 – 59 | 36 260 (17.7) | 15 023 (17.2) | 17 743 (19.1) |
| 60 – 74 | 8 325 (4.1) | 4 330 (5) | 5 372 (5.8) |
| ≥ 75 | 4 896 (2.4) | 6 450 (7.4) | 4 434 (4.8) |
| Unknown |  | 98 (0.1) | 1 326 (1.4) |
| **Sex, n (%)** |  |  |  |
| Male | 109 349 (53.4) | 45 331 (51.9) | 44 774 (48.2) |
| Female | 95 473 (46.6) | 41 656 (47.7) | 47 142 (50.8) |
| Unknown | - | 299 (0.4) | 891 (1) |
| **Diagnosis, n (%)** |  |  |  |
| Respiratory tract infections : |  |  |  |
| Acute upper respiratory tract infections | 53 855 (26.3) | 27 209 (31.2) | 47 046 (50.7) |
| Acute lower respiratory tract infections | 14 003 (6.8) | 16 933 (19.4) | 6 767 (7.3) |
| Influenza | 13 691 (6.7) | 1 465 (1.7) | 1 063 (1.2) |
| Other respiratory tract infections | 28 600 (14) | 1 370 (1.6) | 464 (0.5) |
| Other respiratory acute symptoms | 46 482 (22.7) | 5 361 (6.1) | 1 369 (1.5) |
| Ear infections and mastoiditis | 6 691 (3.8) | 7 374 (8.5) | 6 610 (7.1) |
| Gastrointestinal infections | 45 688 (22.3) | 13 392 (15.3) | 15 546 (16.8) |
| Urinary tract infections (UTIs) |  |  |  |
| Cystitis | 988 (0.5) | 2 309 (2.7) | 730 (0.8) |
| Other complicated/unspecified UTIs | 21 229 (10.4) | 7 035 (8.1) | 5 314 (5.7) |
| Skin infections | 7 036 (3.4) | 5 587 (6.4) | 5 848 (6.3) |
| Fever | 59 936 (29.3) | 7 009 (8) | 3 966 (4.3) |
| **Co-morbidities, n (%)** |  |  |  |
| Another acute infection diagnosis<sup>a</sup> | 70 708 (34.5) | 7 381 (8.5) | 2 162 (2.3) |
| Other co-morbidities<sup>b</sup> | 85 307 (41.7) | 13 112 (15) | 13 188 (14.2) |
| **Having a condition indicating severity, n (%)** |  |  |  |
| Accompanying fever<sup>c</sup> | 58 779 (28.7) | 5 804 (6.6) | 3 813 (4.1) |
| Disease-specific condition<sup>d</sup> | 2 834 (1.4) | 1 097 (1.3) | 325 (0.4) |
| **Type of settings<sup>e</sup>, n (%)** |  |  |  |
| Doctor's clinic | 125 656 (61.3) | 76 967 (88.2) | 38 042 (41) |
| Hospital | 79 166 (38.7) | 5 064 (5.8) | 44 852 (48.3) |
| Other setting/unknown<sup>h</sup> | - | 5 255 (6.0) | 9913 (10.7) |
| **Prescriber' specialisation** |  |  |  |
| General practitioner | 125 358 (61.2) | 14 816 (17) | 50 948 (54.9) |
| Internal medicine | 10 308 (5.0) | 10 724 (12.3) | 5 689 (6.1) |
| Paediatrics | 13 013 (6.4) | 20 597 (23.6) | 14 964 (16.1) |
| Otorhinolaryngology | 7 624 (3.7) | 15 162 (17.4) | 10 184 (11) |
| Pulmonology | 8 265 (4.0) | 10 576 (12.1) | 4 030 (4.3) |
| Nephro-urology | - | 4 830 (5.5) | - |
| Resident Medical Officers | 24 988 (12.2) | - | - |
| Dermatology | 338 (0.7) | 1 473 (1.7) | 1 361 (1.5) |
| Surgery | 2 950 (1.4) | 2 972 (3.4) | 1 808 (2) |
| Others<sup>g</sup> | 10 289 (5.0) | 6 122 (7.0) | 3 003 (3.2) |
| **Prescriber's sex** |  |  |  |
| Male | 188 233 (91.9) | 78 405 (89.8) | 40 011 (43.1) |
| Female | 16 589 (8.1) | 8 881 (10.2) | 52 796 (56.9) |

<sup>a</sup>: Another acute infection diagnosis includes cases with two or more of the acute infections listed above, excluding fever. <sup>b</sup>: Other co-morbidities refer to cases where the patient was diagnosed with two or more conditions, excluding the acute infections listed above. <sup>c</sup>: Accompanying fever was counted for patients with fever alongside any of the acute infection diagnoses listed above, excluding fever itself. <sup>d</sup>: Disease-specific conditions indicating severity include: shortness of breath, difficulty breathing, or chest pain in patients with lower respiratory tract infections; bilateral ear pain, otorrhoea, or ear discharge in patients with otitis; bloody diarrhoea in patients with gastrointestinal infections; blood in urine in patients with urinary tract infections; and swollen lymph nodes in patients with skin infections. <sup>e</sup>: In Pakistan and Egypt, all consultation settings were private. In Indonesia, the majority were private, with only 6.6% (16,840/92,807) conducted in public settings (government hospitals). <sup>h</sup>: other settings included home visits, phone consultations, and consultations where the setting was not specified. <sup>g</sup>: Other specialisation included cardiology, rheumatology, gastroenterology/hepatology, endocrinology, ophthalmology, gynaecology, neurology, psychiatry, dentistry and oncology.
Source: Author analysis of IQVIA Medical Data Index (Medical Index of Pakistan (MIP), Egypt Medical Data Index (EMDI), Indonesia Medical Data Index (IMDI)) for the period 2017-2021, reflecting estimates of real world activity. Copyright IQVIA. All rights reserved.

### Identification of antibiotic treatment

In this study, a consultation was considered to involve antibiotic treatment if at least one systemic antibiotic was prescribed. We determined the antibiotic treatment status for each visit using the WHO AWaRe classification (categorized as Access, Watch, or Reserve) and the WHO Model Essential Medicines List (categorized as EML or Non-EML). For combinations classified as not-recommended or not included by WHO AWaRe, we categorized them based on the antibiotic with the ‘highest’ category within the combination (e.g., a combination of Watch and Access antibiotics was classified as Watch). For single-agent antibiotics not listed in the WHO AWaRe classification, we assigned them to Access, Watch, or Reserve categories by referencing the AWaRe classification of agents within the same chemical groups, typically at the same 4th level of the WHO ATC classification. When multiple antibiotics were used in a visit, the classification of the visit was based on the antibiotic with the highest WHO AWaRe category. Visits with both a non-EML and EML antibiotic were classified as non-EML. We conducted this classification at the consultation level to ensure comparability between consultations in terms of Access, Watch, and Reserve antibiotic use. Our classification of antibiotic treatment was conducted and reviewed by clinicians, infectious disease researchers, and pharmacists. (Appendix 3).

### Statistical analyses

We conducted descriptive analyses of antibiotic prescribing practices, focusing on whether an antibiotic was prescribed and the type of antibiotic prescribed (categorized by WHO AWaRe and WHO EML) for health visits related to common infections in each country.

We analysed the determinants of the decision to prescribe an antibiotic or not (yes/no), and Watch antibiotic prescribing compared to Access antibiotics and a no-antibiotic strategy (Watch/Access/no antibiotic). We used mixed-effects multinomial regression models (mclogit package in R) to estimate odds ratios (ORs) and 95% confidence intervals (CIs). Variables included in the model were identified through a literature review of similar studies, considering the availability and accuracy of data on potential covariates in our dataset. Fixed effects included patient-specific factors (age; sex; clinical diagnosis; co-diagnosis with other conditions; presence of accompanying clinical signs indicating severe infection) and prescriber-specific factors (sex of the prescriber, prescriber’s specialty, and the type of clinical setting where the consultation occurred). Prescriber ID was included as a random effect to account for clustering by different health settings.

Statistical analysis was conducted using R version 4.4.1.

### Ethics

This study is a secondary data analysis using non-identifiable data from IQVIA’s Medical Data Index and, as such, does not require ethics approval.

### Patient and public involvement

Patients and the public were not involved in this research study.

## RESULTS

Our analysis included a total of 384,975 consultation records from primary care and outpatient visits for acute common infections, with 204,882 records from Pakistan, 87,286 from Egypt, and 92,807 from Indonesia. The median age of the patients was similar between the countries, ranging from 25 years [IQR 10-42] (Pakistan) to 27 years [IQR 10-42] (Indonesia). The most common infectious diagnoses were acute upper RTIs (Pakistan: 26.3%; Egypt: 31.3% and Indonesia: 50.7%) and GIs (Pakistan: 22.3%; Egypt: 13.4% and Indonesia: 16.8%).

Fever was more often documented in recorded consultations in Pakistan (29.3%) than in Egypt (8.0%) and Indonesia (4.3%). In Pakistan and Egypt, the majority of records came from doctors’ clinics (61.4% and 88.2%, respectively) while in Indonesia, the distribution of records were more balanced across different types of settings. In each country, 80% of prescriptions were issued by prescribers specialising in one of the following fields: general practice or internal medicine, paediatrics, otorhinolaryngology, or pulmonology (Table 1).

### Antibiotic prescribing by country

Antibiotic treatment was prescribed in 82% of consultations in Pakistan, 81.2% in Egypt and 69.1% in Indonesia. Watch antibiotics accounted for 70.2% of all antibiotic prescriptions in Pakistan, 52.9% in Egypt and 53.6% in Indonesia. Access antibiotics were used less frequently (Pakistan: 24.2%, Egypt: 45.9% and Indonesia: 46.3%) and Reserve antibiotics were rarely used (Pakistan: 0.5%, Egypt: 1.2% and Indonesia: 0.1%). The use of non-EML antibiotics was also high, accounting for 26.8% of all antibiotic prescriptions in Pakistan, 39.9% in Egypt and 33.0% in Indonesia (Table 2).

**Table 2.**
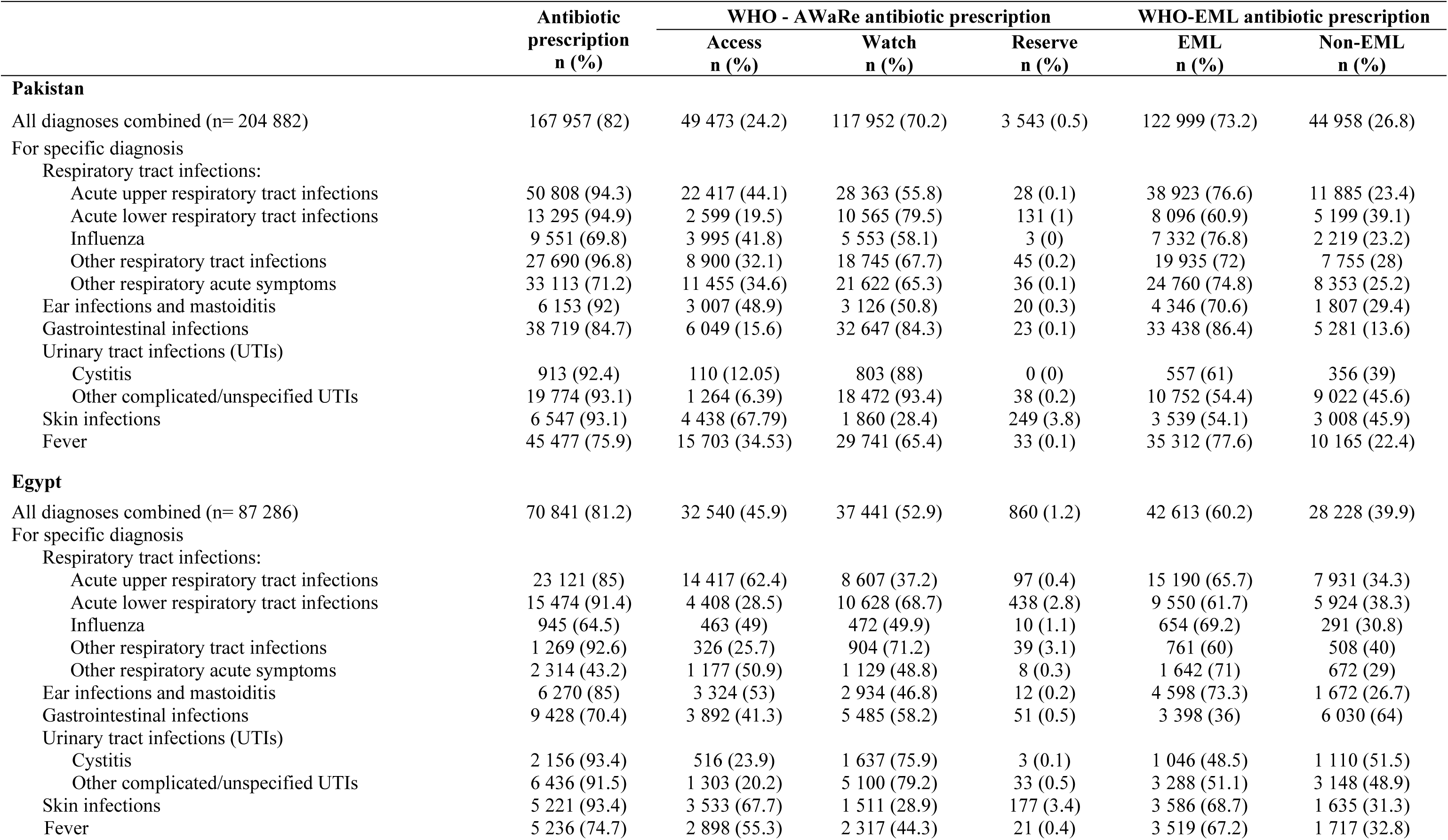
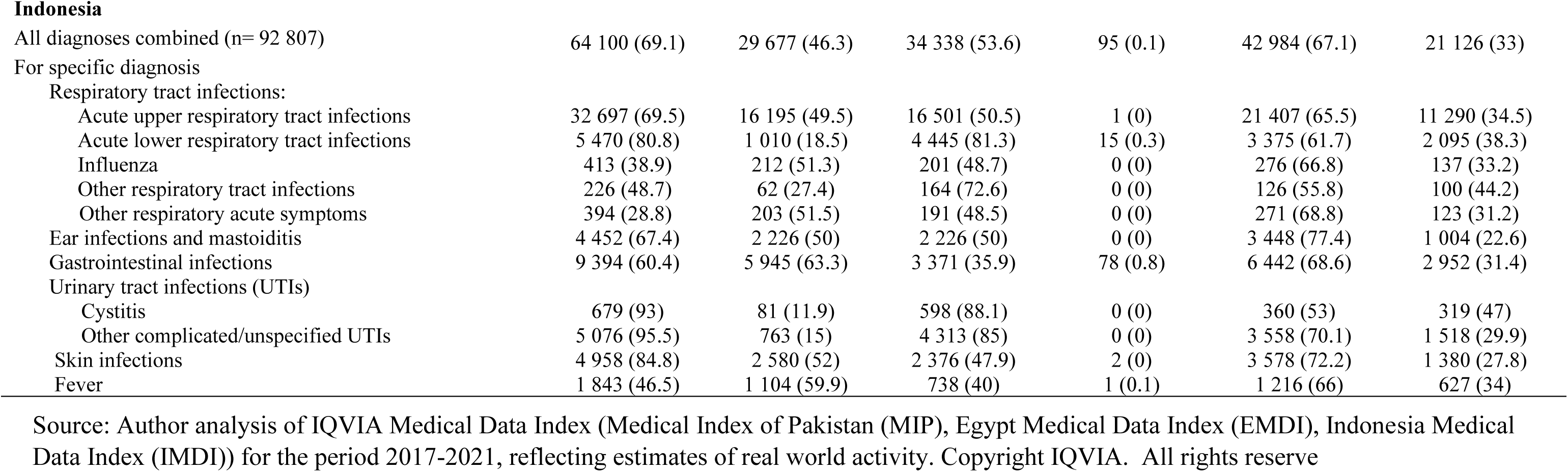
Antibiotic prescribing patterns by infectious group and by country.

|  | Antibiotic<br>prescription<br>n (%) | WHO - AWaRe antibiotic prescription |  |  | WHO-EML antibiotic prescription |  |
| --- | --- | --- | --- | --- | --- | --- |
|  |  | Access<br>n (%) | Watch<br>n (%) | Reserve<br>n (%) | EML<br>n (%) | Non-EML<br>n (%) |
| **Pakistan** |  |  |  |  |  |  |
| All diagnoses combined (n= 204 882) | 167 957 (82) | 49 473 (24.2) | 117 952 (70.2) | 3 543 (0.5) | 122 999 (73.2) | 44 958 (26.8) |
| For specific diagnosis |  |  |  |  |  |  |
| Respiratory tract infections: |  |  |  |  |  |  |
| Acute upper respiratory tract infections | 50 808 (94.3) | 22 417 (44.1) | 28 363 (55.8) | 28 (0.1) | 38 923 (76.6) | 11 885 (23.4) |
| Acute lower respiratory tract infections | 13 295 (94.9) | 2 599 (19.5) | 10 565 (79.5) | 131 (1) | 8 096 (60.9) | 5 199 (39.1) |
| Influenza | 9 551 (69.8) | 3 995 (41.8) | 5 553 (58.1) | 3 (0) | 7 332 (76.8) | 2 219 (23.2) |
| Other respiratory tract infections | 27 690 (96.8) | 8 900 (32.1) | 18 745 (67.7) | 45 (0.2) | 19 935 (72) | 7 755 (28) |
| Other respiratory acute symptoms | 33 113 (71.2) | 11 455 (34.6) | 21 622 (65.3) | 36 (0.1) | 24 760 (74.8) | 8 353 (25.2) |
| Ear infections and mastoiditis | 6 153 (92) | 3 007 (48.9) | 3 126 (50.8) | 20 (0.3) | 4 346 (70.6) | 1 807 (29.4) |
| Gastrointestinal infections | 38 719 (84.7) | 6 049 (15.6) | 32 647 (84.3) | 23 (0.1) | 33 438 (86.4) | 5 281 (13.6) |
| Urinary tract infections (UTIs) |  |  |  |  |  |  |
| Cystitis | 913 (92.4) | 110 (12.05) | 803 (88) | 0 (0) | 557 (61) | 356 (39) |
| Other complicated/unspecified UTIs | 19 774 (93.1) | 1 264 (6.39) | 18 472 (93.4) | 38 (0.2) | 10 752 (54.4) | 9 022 (45.6) |
| Skin infections | 6 547 (93.1) | 4 438 (67.79) | 1 860 (28.4) | 249 (3.8) | 3 539 (54.1) | 3 008 (45.9) |
| Fever | 45 477 (75.9) | 15 703 (34.53) | 29 741 (65.4) | 33 (0.1) | 35 312 (77.6) | 10 165 (22.4) |
| **Egypt** |  |  |  |  |  |  |
| All diagnoses combined (n= 87 286) | 70 841 (81.2) | 32 540 (45.9) | 37 441 (52.9) | 860 (1.2) | 42 613 (60.2) | 28 228 (39.9) |
| For specific diagnosis |  |  |  |  |  |  |
| Respiratory tract infections: |  |  |  |  |  |  |
| Acute upper respiratory tract infections | 23 121 (85) | 14 417 (62.4) | 8 607 (37.2) | 97 (0.4) | 15 190 (65.7) | 7 931 (34.3) |
| Acute lower respiratory tract infections | 15 474 (91.4) | 4 408 (28.5) | 10 628 (68.7) | 438 (2.8) | 9 550 (61.7) | 5 924 (38.3) |
| Influenza | 945 (64.5) | 463 (49) | 472 (49.9) | 10 (1.1) | 654 (69.2) | 291 (30.8) |
| Other respiratory tract infections | 1 269 (92.6) | 326 (25.7) | 904 (71.2) | 39 (3.1) | 761 (60) | 508 (40) |
| Other respiratory acute symptoms | 2 314 (43.2) | 1 177 (50.9) | 1 129 (48.8) | 8 (0.3) | 1 642 (71) | 672 (29) |
| Ear infections and mastoiditis | 6 270 (85) | 3 324 (53) | 2 934 (46.8) | 12 (0.2) | 4 598 (73.3) | 1 672 (26.7) |
| Gastrointestinal infections | 9 428 (70.4) | 3 892 (41.3) | 5 485 (58.2) | 51 (0.5) | 3 398 (36) | 6 030 (64) |
| Urinary tract infections (UTIs) |  |  |  |  |  |  |
| Cystitis | 2 156 (93.4) | 516 (23.9) | 1 637 (75.9) | 3 (0.1) | 1 046 (48.5) | 1 110 (51.5) |
| Other complicated/unspecified UTIs | 6 436 (91.5) | 1 303 (20.2) | 5 100 (79.2) | 33 (0.5) | 3 288 (51.1) | 3 148 (48.9) |
| Skin infections | 5 221 (93.4) | 3 533 (67.7) | 1 511 (28.9) | 177 (3.4) | 3 586 (68.7) | 1 635 (31.3) |
| Fever | 5 236 (74.7) | 2 898 (55.3) | 2 317 (44.3) | 21 (0.4) | 3 519 (67.2) | 1 717 (32.8) |
| Indonesia |  |  |  |  |  |  |
| All diagnoses combined (n= 92 807) | 64 100 (69.1) | 29 677 (46.3) | 34 338 (53.6) | 95 (0.1) | 42 984 (67.1) | 21 126 (33) |
| For specific diagnosis |  |  |  |  |  |  |
| Respiratory tract infections: |  |  |  |  |  |  |
| Acute upper respiratory tract infections | 32 697 (69.5) | 16 195 (49.5) | 16 501 (50.5) | 1 (0) | 21 407 (65.5) | 11 290 (34.5) |
| Acute lower respiratory tract infections | 5 470 (80.8) | 1 010 (18.5) | 4 445 (81.3) | 15 (0.3) | 3 375 (61.7) | 2 095 (38.3) |
| Influenza | 413 (38.9) | 212 (51.3) | 201 (48.7) | 0 (0) | 276 (66.8) | 137 (33.2) |
| Other respiratory tract infections | 226 (48.7) | 62 (27.4) | 164 (72.6) | 0 (0) | 126 (55.8) | 100 (44.2) |
| Other respiratory acute symptoms | 394 (28.8) | 203 (51.5) | 191 (48.5) | 0 (0) | 271 (68.8) | 123 (31.2) |
| Ear infections and mastoiditis | 4 452 (67.4) | 2 226 (50) | 2 226 (50) | 0 (0) | 3 448 (77.4) | 1 004 (22.6) |
| Gastrointestinal infections | 9 394 (60.4) | 5 945 (63.3) | 3 371 (35.9) | 78 (0.8) | 6 442 (68.6) | 2 952 (31.4) |
| Urinary tract infections (UTIs) |  |  |  |  |  |  |
| Cystitis | 679 (93) | 81 (11.9) | 598 (88.1) | 0 (0) | 360 (53) | 319 (47) |
| Other complicated/unspecified UTIs | 5 076 (95.5) | 763 (15) | 4 313 (85) | 0 (0) | 3 558 (70.1) | 1 518 (29.9) |
| Skin infections | 4 958 (84.8) | 2 580 (52) | 2 376 (47.9) | 2 (0) | 3 578 (72.2) | 1 380 (27.8) |
| Fever | 1 843 (46.5) | 1 104 (59.9) | 738 (40) | 1 (0.1) | 1 216 (66) | 627 (34) |

Source: Author analysis of IQVIA Medical Data Index (Medical Index of Pakistan (MIP), Egypt Medical Data Index (EMDI), Indonesia Medical Data Index (IMDI)) for the period 2017-2021, reflecting estimates of real world activity. Copyright IQVIA. All rights reserve

### Antibiotic prescribing by infection type

When the data were stratified by the 11 infection diagnoses, we found that antibiotic treatment was prescribed for over 80% of health consultations for most infection types in Pakistan (except influenza at 69.8% and other respiratory acute symptoms at 71.2%) and Egypt (except influenza at 64.5% and other acute respiratory symptoms at 43.2%). In Indonesia, where the overall antibiotic treatment proportion was slightly lower (around 70% compared to 80% in Pakistan and Egypt), common infections like acute upper RTIs and GIs were still frequently treated with antibiotics (69.5% and 60.4%, respectively). Watch antibiotics were used in over 50% of antibiotic prescriptions for 10 out of 11 infections in Pakistan, 6 out of 11 in Egypt, and 6 out of 11 in Indonesia (Table 2).

In all three countries, the most common pharmacological groups of antibiotics were those related to Watch antibiotics, including second/third-generation cephalosporins (mostly third-generation), quinolones and macrolides. Together, these three antibiotic classes accounted for 50%-75% of antibiotic prescriptions for most infections. The use of penicillins, such as amoxicillin, which are generally recommended as first-line Access antibiotics for most common infections^17^, was very low (less than 5%) for most infections in Pakistan and Egypt. The use of penicillins was more common in Indonesia, but remained below 25% of total antibiotic prescriptions for most infections (10/11, except for influenza, for which antibiotics are not recommended) (Figure 1 and Appendix 4).

**Figure 1.**
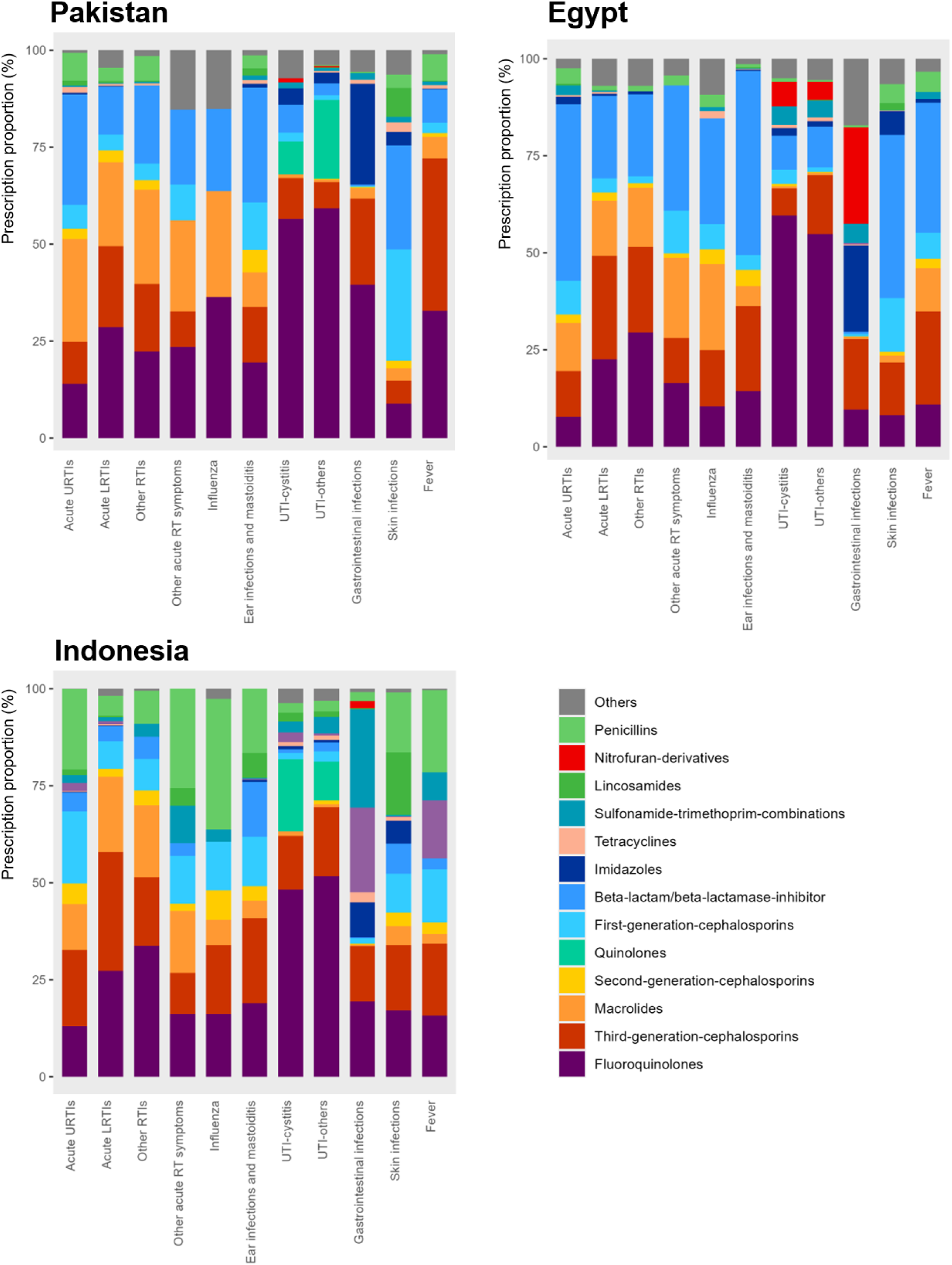
Distribution of antibiotic classes, by infectious group and by country. The pharmacological grouping follows the WHO AWaRe classification. https://www.who.int/publications/i/item/WHO-MHP-HPS-EML-2023.04 Source: Author analysis of IQVIA Medical Data Index (Medical Index of Pakistan (MIP), Egypt Medical Data Index (EMDI), Indonesia Medical Data Index (IMDI)) for the period 2017-2021, reflecting estimates of real world activity. Copyright IQVIA. All rights reserved.

### Antibiotic prescribing by prescriber’s speciality

We observed consistently high antibiotic prescribing for common infections across most specialties in the studied countries—exceeding 75% in Pakistan and Egypt, and over 70% in Indonesia (Figure 2A and Appendix 5). When grouping the data by prescriber the median proportion of patients receiving any antibiotic prescription was high (above 75% in Pakistan and Egypt and above 60% in Indonesia) and the median proportions of prescribing Watch antibiotics were generally above 50%. (Figure 2B1-2) Variation in the proportion of consultations receiving any antibiotic or Watch antibiotic within the same specialty was least pronounced in Pakistan, higher in Egypt and most pronounced in Indonesia, as indicated by a lower spread in the antibiotic prescribing proportions among prescribers within the same specialty. For example, the interquartile range of the proportion of consultations receiving Watch antibiotics in general practices in Pakistan was 13.8%, compared to 28.2% in Egypt, and 33.6% in Indonesia. This pattern was consistent across other medical specialties. (Figure 2B1-2 and Appendix 6)

**Figure 2.**
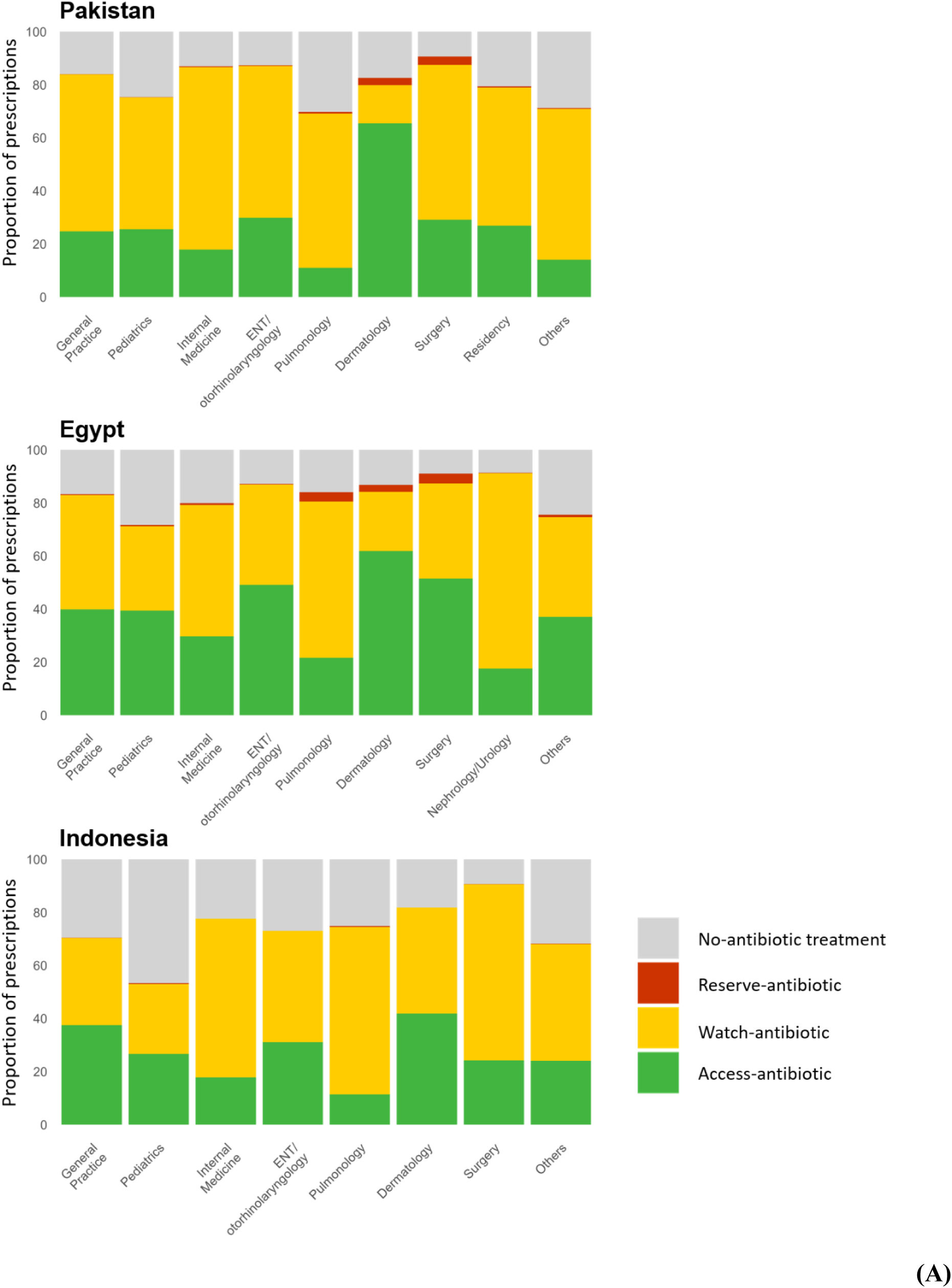
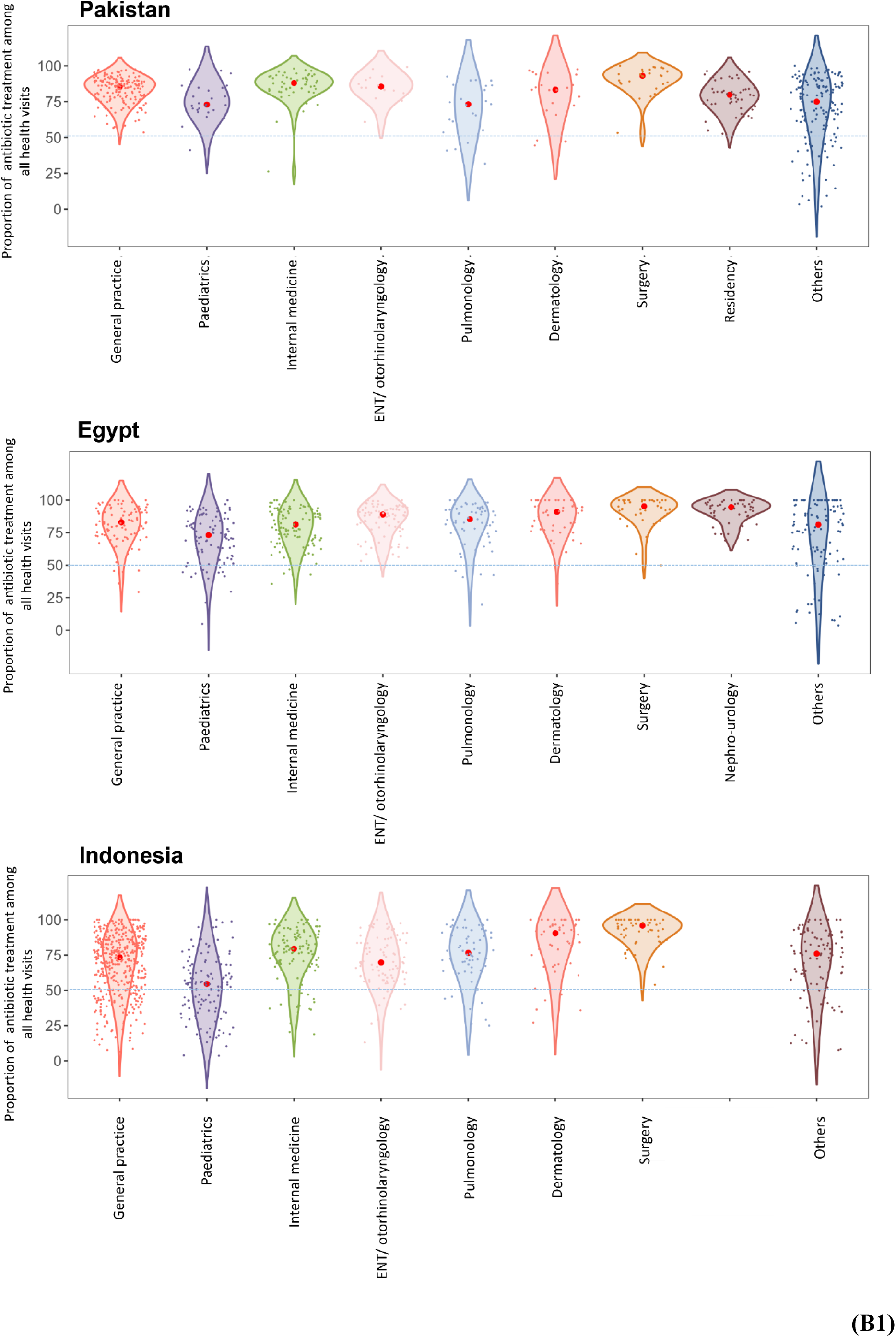
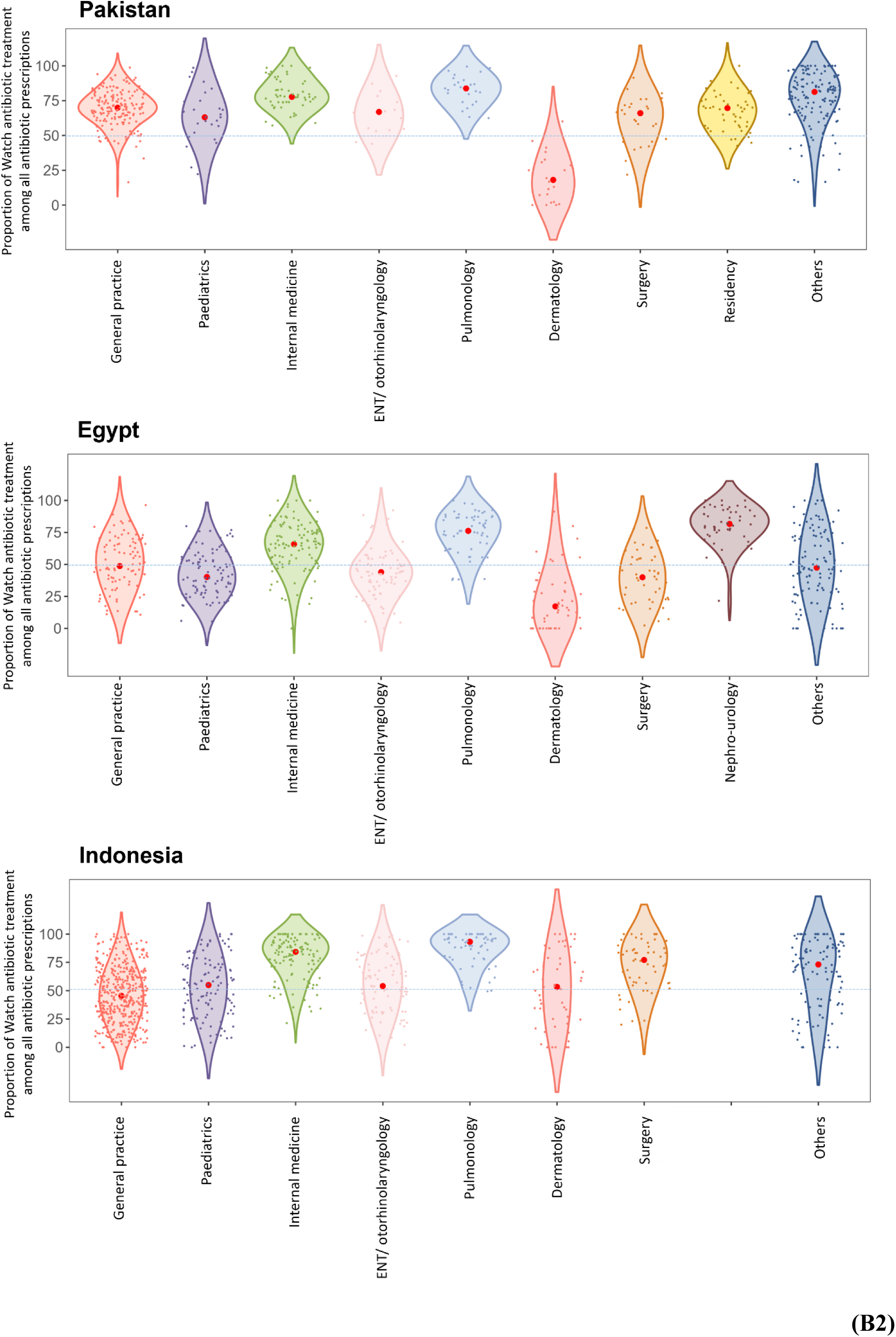
(A) Distributional proportions of WHO AWaRe antibiotic prescribing by prescriber’s speciality and by country. (B1) Variability in proportions of antibiotic prescriptions among doctors within and between specialities. (B2) Variability in proportions of Watch-antibiotic prescriptions relative to all antibiotic prescriptions among doctors within and between specialities. In Figures 2B1 and 2B2, the x-axis represents the specialities of prescribers who participated in the study in each country, while the y-axis represents the proportion of antibiotic prescriptions (2B1) and Watch-antibiotic prescriptions (2B2) by individual doctors, expressed as percentages ranging from 0% to 100%. Each dot corresponds to a specific doctor’s prescribing proportion. Due to the kernel density estimation used in the violin plot, the visualization may extend beyond this range, even though actual data points are confined within 0% to 100%. Source: Author analysis of IQVIA Medical Data Index (Medical Index of Pakistan (MIP), Egypt Medical Data Index (EMDI), Indonesia Medical Data Index (IMDI)) for the period 2017-2021, reflecting estimates of real world activity. Copyright IQVIA. All rights reserved.

### Determinants for antibiotic prescribing decisions

In all recorded consultations across the all three countries, increasing patient age was associated with a gradual reduction in the odds of prescribing any antibiotic, most notably in Egypt (Figure 3A, 3B and 3C). Among consultations involving antibiotic treatment, consultations with patients aged 40-59 had higher odds of receiving Watch antibiotics over Access antibiotics (referred to as odds of receiving Watch antibiotics from now on) compared to those aged 25– 39, while patients under 18 and over 75 had lower odds.

**Figure 3.**
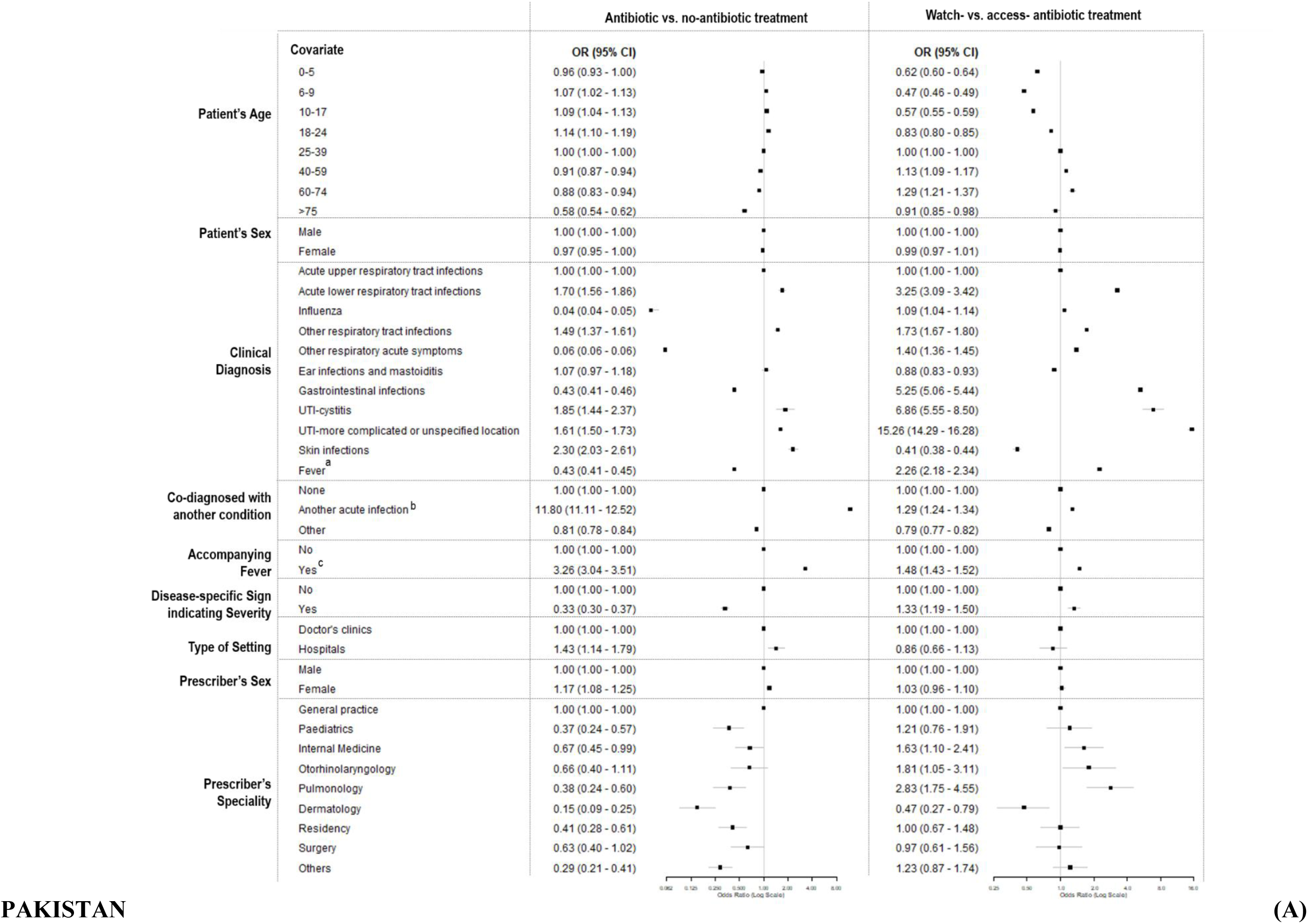
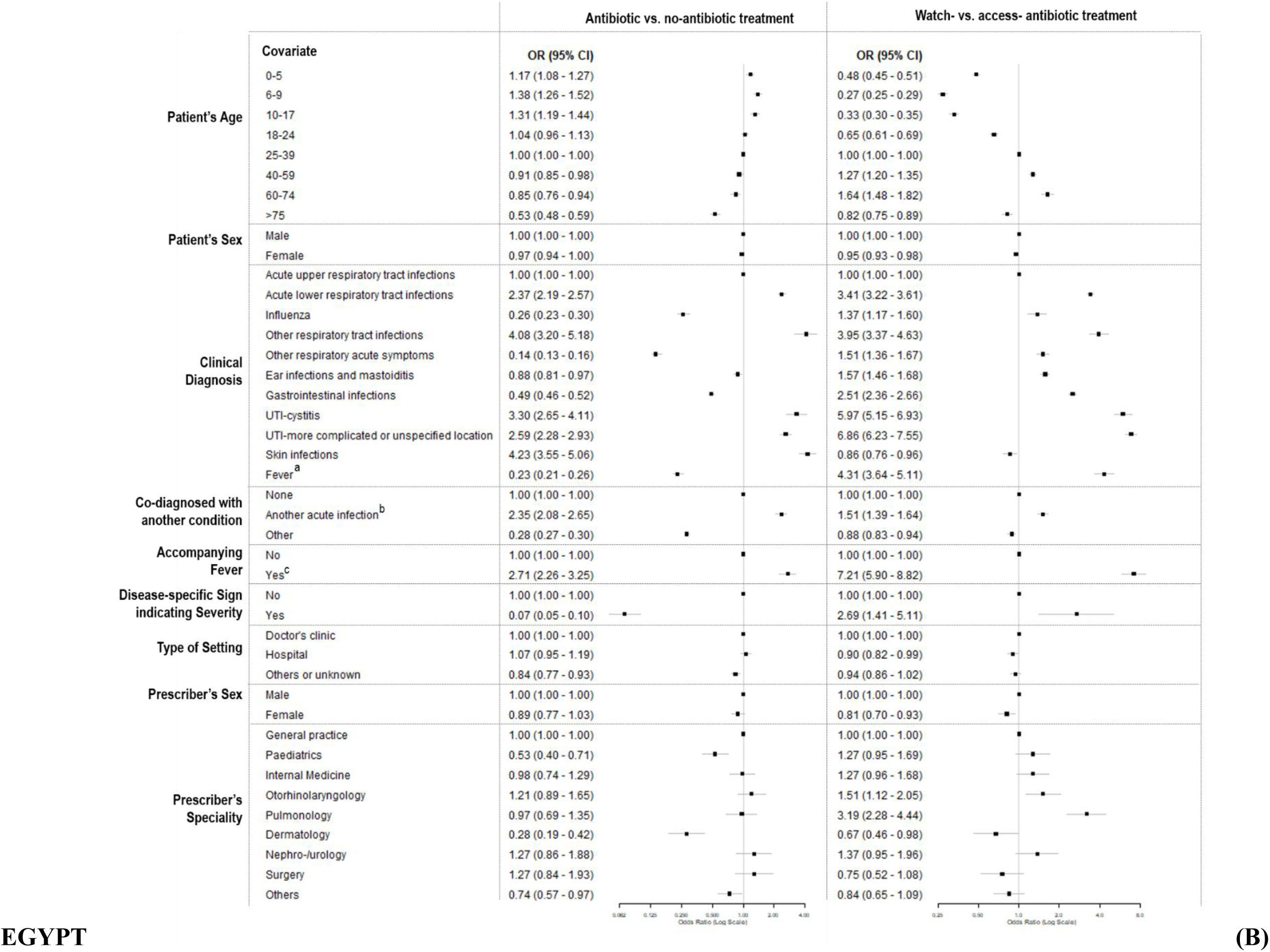
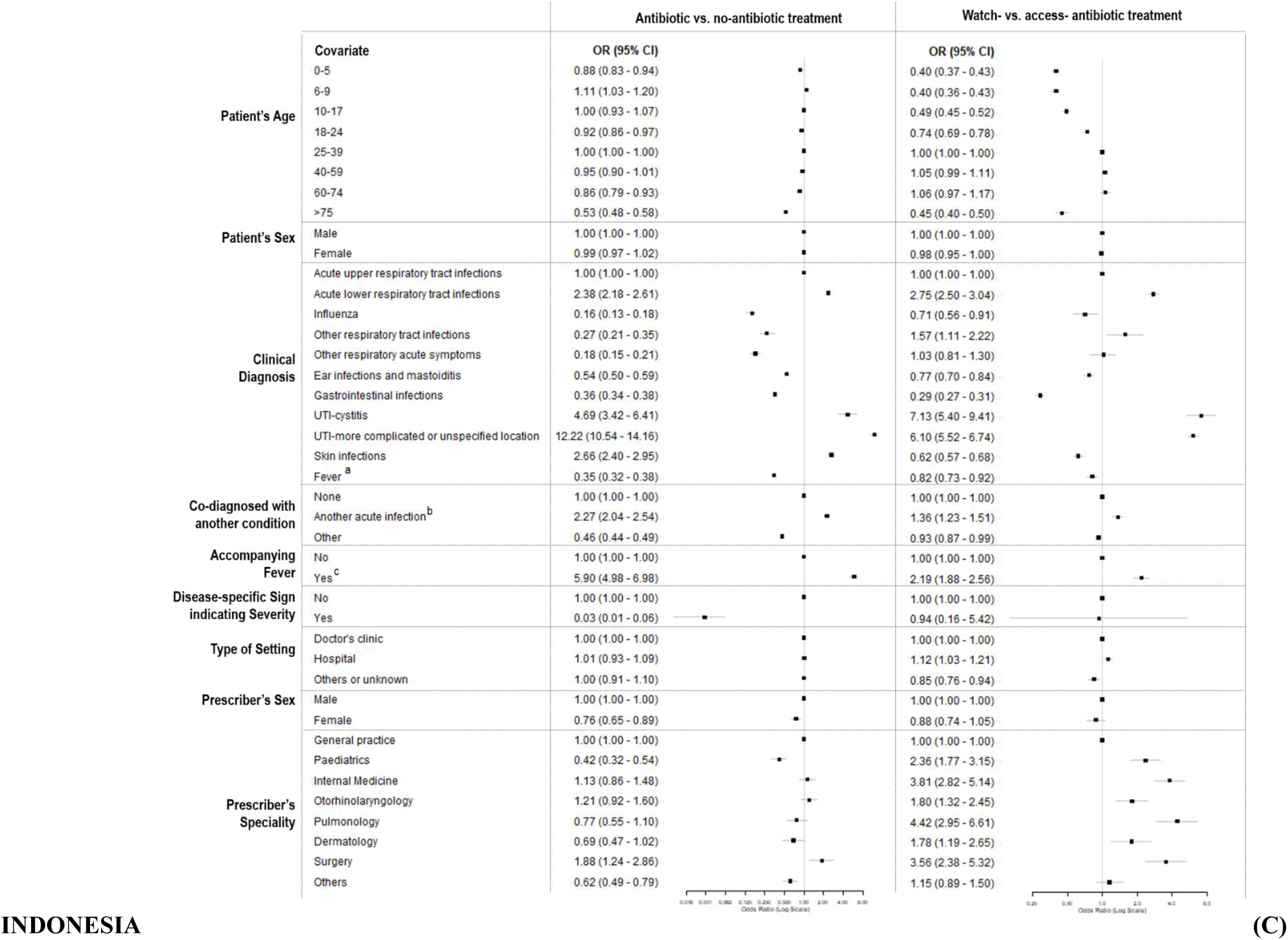
Factors associated with antibiotic prescribing decisions, including whether antibiotics were given and the choice between Watch-antibiotics and Access-antibiotics, for patients in Pakistan (3A), Egypt (3B), and Indonesia (3C). a: cases with single diagnosis of fever. b: cases with more than one diagnosis of acute infections. c: cases with fever accompanying an acute infection. Source: Author analysis of IQVIA Medical Data Index (Medical Index of Pakistan (MIP), Egypt Medical Data Index (EMDI), Indonesia Medical Data Index (IMDI)) for the period 2017-2021, reflecting estimates of real world activity. Copyright IQVIA. All rights reserved.

When compared to consultations for upper RTIs, those for lower RTIs, cystitis, and other UTIs had the highest odds of including any antibiotics as well as Watch antibiotics. The adjusted odds ratios (aORs) for prescribing any antibiotics were: lower RTIs (Pakistan: 1.70, 95% CI 1.56-1.86; Egypt: 2.37, 2.19-2.57; Indonesia: 2.38, 2.18-2.61), cystitis (Pakistan: 1.85, 1.44-2.37; Egypt: 3.30, 2.65-4.11; Indonesia: 4.69, 3.42-6.41), and other UTIs (Pakistan: 1.61, 1.50-1.73, Egypt: 2.59, 2.28-2.93; Indonesia: 12.22, 10.54-14.16). The aORs for receiving Watch antibiotics were: lower RTIs (Pakistan: 3.25, 3.09-3.42; Egypt: 3.41, 3.22-3.61; Indonesia: 2.75, 2.50-3.04), cystitis (Pakistan: 6.86, 5.55-8.50; Egypt: 5.97, 5.15-6.93; Indonesia: 7.13, 5.40-9.41), and other UTIs (Pakistan: 15.26, 14.29-16.28; Egypt: 6.86, 6.23-7.55; Indonesia: 6.10, 5.52-6.74). Several other conditions were associated with decreased odds of prescribing any antibiotics but elevated odds of prescribing Watch antibiotics, although this was not entirely consistent across all three countries. For example, in Pakistan, the aORs for prescribing any antibiotics in consultations for patients with GIs was 0.43 (0.41-0.46), while that for prescribing Watch antibiotics was 5.25 (5.06-5.44). While similar patterns were observed in Egypt, in Indonesia, patients with GIs were linked to decreased odds of prescribing both any (0.36, 0.34-0.38) and Watch (0.29, 0.27-0.31) antibiotics.

Compared to consultations for a single infection diagnosis, those with multiple acute infection diagnoses had elevated odds of prescribing any antibiotics (Pakistan: 11.80, 11.11–12.52; Egypt: 2.35, 2.08–2.65; Indonesia: 2.27, 2.04–2.95) or Watch antibiotics (Pakistan: 1.29, 1.24–1.34; Egypt: 1.51, 1.39–1.64; Indonesia: 1.36, 1.23–1.51). In contrast, consolations for patients co-diagnosed with conditions outside of acute infections generally had reduced odds of prescribing antibiotics (e.g., Pakistan: 0.81, 0.78–0.84 for any antibiotic prescription and 0.79, 0.77–0.82 for Watch antibiotic prescriptions).

Consultations for patients with an infection diagnosis and recorded fever had higher odds of prescribing any antibiotics (Pakistan: 3.26, 3.04–3.51; Egypt: 2.35, 2.08–2.65; Indonesia: 2.27, 2.04–2.95) or Watch antibiotics (Pakistan: 1.48, 1.43–1.52; Egypt: 2.35, 2.08–2.65; Indonesia: 2.27, 2.04–2.95) compared to those with an infectious diagnosis but no fever. In Pakistan and Egypt, consultations for patients with disease-specific signs indicating severity (e.g., chest pain in patients with lower respiratory tract symptoms) had lower odds of prescribing antibiotics at the primary care level (Pakistan: 0.33, 0.30–0.37; Egypt: 0.07, 0.05–0.10), but if an antibiotic was prescribed, they had higher odds of prescribing a Watch antibiotic (Pakistan: 1.33, 1.19– 1.50; Egypt: 2.69, 1.41–5.11). A similar trend was observed in Indonesia for any antibiotic prescription (0.03, 0.01–0.06), but there was no difference in the odds of prescribing Watch antibiotics (0.94, 0.16–5.42).

There was no substantial difference in the odds of prescribing either any antibiotics or Watch antibiotics between consultations at doctors’ clinics and hospitals across the three countries except that hospital consultations in Pakistan had elevated odds of antibiotic treatments (1.43, 1.14-1.79). Consultations by female doctors in Indonesia had lower odds of including antibiotics (0.76, 0.65–0.89) and in Egypt had lower odds of including Watch antibiotics (0.81, 0.70–0.93). In contrast, consultations by female doctors in Pakistan were associated with higher odds of including antibiotics (1.17, 1.08–1.25). Compared to consultations by general practitioners, consultations by respiratory-related specialists in Pakistan (otorhinolaryngologists: 1.81, 1.05–3.11; pulmonologists: 2.83, 1.75–4.55) and Egypt (otorhinolaryngologists: 1.51, 1.12–2.05; pulmonologists: 3.19, 2.28–4.44) and with most specialists in Indonesia (paediatricians: 2.36, 1.77–3.15; internal medicine doctors: 3.81, 2.83–5.14; otorhinolaryngologists: 1.80, 1.32–2.45; pulmonologists: 4.42, 2.95–6.61; dermatologists: 1.78, 1.19–2.65; surgeons: 3.56, 2.38–5.32) had higher odds of prescribing Watch antibiotics.

## DISCUSSION

To our knowledge this is the largest patient level analysis of primary care and outpatient antibiotic prescribing in the private sector across multiple low- and middle-income countries. We observed antibiotic prescribing proportions as high as 70 to 80% of patient visits for common acute infections, including those that can generally be safely managed without antibiotics or for which antibiotics are not indicated, such as upper RTIs and viral influenza (see Appendix 7).^16^ Use of Watch antibiotics was common, despite the AWaRe book guidance indicating that most infections reported here should be treated with Access antibiotics or no antibiotics care.^14^

Some of the common Watch antibiotic use may be due to the empiric treatment of suspected typhoid fever, which has a high incidence in Pakistan and Indonesia, and to a much lesser extent in Egypt.^18^ Watch antibiotics recommended for the treatment of uncomplicated typhoid fever in local guidelines in Pakistan were commonly used for patients with fever in our study (Appendix 4). Typhoid fever can be difficult to diagnose without diagnostic tests that are generally not widely available in LMICs, due to symptom overlap with infection types that often do not require antibiotics, such as signs of upper RTIs.^19^ Introduction of typhoid vaccines into routine immunization programs could remove this barrier to reductions in antibiotic use.^20^ To effectively reduce antibiotic prescribing in high-endemic countries the vaccination program should ideally be complemented by updated local prescribing guidance, training of prescribers, and programs raising the awareness among the general public to appropriately reflect changed pre-test probabilities.^21^ Introducing a typhoid vaccination program in the capital of Zimbabwe alone was, though, not associated with a reduction in antibiotic use the first year after the program started.^21,22^

Patients presenting with fever were more likely to receive antibiotics, including Watch antibiotics. Fever is an important component of the FeverPAIN and Centor scores which are commonly used to guide whether (Access) antibiotics are needed for patients presenting with a sore throat.^23^ Furthermore, when UTIs are accompanied by fever the patient may have pyelonephritis for which the AWaRe guidance suggests a Watch antibiotic.

Reduced odds of being prescribed antibiotics in doctor visits for infections concurrently diagnosed with another non-infectious condition (compared to visits with a single infectious diagnosis) and for visits with disease-specific signs indicating severity (compared to those without such signs) could be explained by several mechanisms. First, those patients could be immediately admitted to hospital for antibiotic treatment. Second, some co-diagnosed conditions or signs indicating severity are not specific for acute infections and might be attributed to underlying chronic conditions of the patients. For example, patients with acute cough and chest pain or shortness of breath due to underlying cardiovascular disease, or those with gastroesophageal reflux disease presenting with acute cough, may have a lower likelihood of being treated with antibiotics compared to patients with a single specific infectious diagnosis.^24,25^

Antibiotic prescribing decisions were not equal among all specialists, with adjusted odds of receiving Watch antibiotics being higher among patients treated by respiratory-related specialists in Pakistan and Egypt, and by most specialists in Indonesia. In LMICs, where specialists are involved in both primary and secondary care through their dual practice (i.e. having a private clinic as well as a public facility role), they may drive the increased use of Watch antibiotics in the community by applying hospital-level prescribing practices to patients with minor infections.^26,27^ Beyond patient- and prescriber-related factors, pharmaceutical suppliers may also play a role in Watch antibiotic overuse. The greatest number of Watch antibiotic products and suppliers was found in Pakistan (Appendix 8), the country with the highest proportion of Watch antibiotic prescriptions. This may increase pressure on prescribers to issue Watch antibiotics, although it could also reflect prescriber preferences within Pakistan’s private sector. Over-prescription of antibiotics for common infections in Pakistan, Egypt, and Indonesia has been documented previously.^24–26^ Overall, these studies reported either lower antibiotic prescribing proportions or a less predominant use of Watch antibiotics compared to our findings. However, most of the previous studies were conducted in public healthcare settings. In these settings, antibiotic prescribing is regulated by health authorities and public reimbursement agencies, which may reduce antibiotic over-prescription. However, private clinics and hospitals are common—if not the most common—points of care for minor illnesses in these countries.^28–30^ With fewer regulatory restrictions, prescribing in private settings is largely influenced by prescriber preferences and patient demand, often driving antibiotic overuse. While our study focuses mainly on private settings, we found that outpatient consultations in private hospitals had higher odds of receiving Watch-over Access-antibiotics than those in public hospitals in Indonesia (Appendix 9).

Among the three countries studied, Pakistan showed the highest antibiotic prescribing proportion, the most frequent use of Watch antibiotics, and the lowest variability on total antibiotic and Watch antibiotic prescribing proportions among prescribers within and across specialties. This low variability may indicate a strong social norm regarding antibiotic use among prescribers, which is a major challenge when aiming to change prescribing behaviours.^31^ Previous studies have also emphasized the overuse and inappropriate use of antibiotics in Pakistan, alongside a culture of self-medication and purchasing antibiotics without prescriptions, all contributing to the high AMR burden in Pakistan.^2^

An important strength of this study is that it is one of the largest studies on antibiotic prescribing in LMICs, covering a wide range of infections and medical specialties while minimizing recall bias and missing information on potential patient and prescriber-level characteristics through analysing data prospectively collected independent of antibiotic prescribing decisions. Our study is the largest to examine prescribing practices in private settings in LMICs. We also provided information on variability in prescribing practice among prescribers and identified which factors may be driving antibiotic prescribing decisions.

This study has several limitations. The study sample may have limited representativeness due to a lack of coverage in certain regions in each country included, and its focus on the private sector. Doctors were required to report health records within only one week per quarter/semester during 2017-2020, which might be considered too short to fully represent their routine prescribing practices. Although the study protocol aimed to collect data on all health consultations, regardless of whether a prescription was issued, no visits without a prescription were recorded. However, this may reflect high patient demand for out-of-pocket services in private settings, where it is rare for patients to leave without a drug (prescription). Patients with common infections in our sample may have been more severe, potentially leading to an overestimation of antibiotic overuse, as those with milder symptoms may go to drug stores instead of paying out-of-pocket for private care. It is possible that only prescribers confident in their prescribing practices agreed to participate in the surveys, causing an underestimation of the actual extent of antibiotic overuse. The self-reported prescription data may have reduced but not fully eliminated social desirability bias. Doctors might still report appropriate prescribing based on their perception, potentially underestimating antibiotic overprescribing.

In conclusion, very high levels of antibiotic prescribing and predominant use of Watch antibiotics for common acute infections were observed across most infection types and prescribers in Pakistan, Egypt, and Indonesia.

### Contributors

NN, KP, MS and AC conceptualised the project. NN and KP developed the methods. NN conducted the formal analysis under KP’s supervision, with support from MT. SM and PS accessed the data, ran the codes prepared by NN, and verified the data. NN, KP, MS, and PS reviewed and validated the results. NN drafted the manuscript under the supervision of KP and MS, with all authors contributing to revisions and edits. PS and AC managed the project, while KP and MS secured funding and provided overall supervision. All authors had final responsibility for the decision to submit for publication.

## Data sharing

The statements, findings, conclusions, views and opinions contained and expressed in this research article are based in part on data obtained under licence from the following IQVIA information service(s): namely IQVIA’s proprietary Medical Data Index (Medical Index of Pakistan (MIP), Egypt Medical Data Index (EMDI) and Indonesia Medical Data Index (IMDI)), available on a confidential basis by subscription from IQVIA. Copyright IQVIA. All rights reserved. IQVIA market research information reflects estimates of real world activity and should be treated accordingly.

## Data Availability

The statements, findings, conclusions, views and opinions contained and expressed in this research article are based in part on data obtained under licence from the following IQVIA information service(s): namely IQVIAs proprietary Medical Data Index (Medical Index of Pakistan (MIP), Egypt Medical Data Index (EMDI) and Indonesia Medical Data Index (IMDI)), available on a confidential basis by subscription from IQVIA. Copyright IQVIA. All rights reserved. IQVIA market research information reflects estimates of real world activity and should be treated accordingly.

## Acknowledgement

The statements, findings, conclusions, views, and opinions contained and expressed herein are not necessarily those of IQVIA.

---

**Appendix 1.** The total number of doctors submitting records of health consultations (for any visits or visits related to common conditions) and the total number of records of health consultations submitted (for any visits or common conditions) were recorded per quarter or term in each country: A) Pakistan, B) Egypt, C) Indonesia

Source: Author analysis of IQVIA Medical Data Index (Medical Index of Pakistan (MIP), Egypt Medical Data Index (EMDI), Indonesia Medical Data Index (IMDI)) for the period 2017-2021, reflecting estimates of real world activity. Copyright IQVIA. All rights reserved.

A. PAKISTAN
| Region | Quarter/<br>Term | Year | Total number of<br>doctors submitting<br>records of health<br>consultations for<br>any condition | Number of<br>records of<br>health<br>consultations<br>submitted | Number of doctors<br>submitting records<br>of health<br>consultations for<br>common infection-related visits | Number of<br>records of<br>health<br>consultations<br>for common<br>infection-related visits |
| --- | --- | --- | --- | --- | --- | --- |
| Baluchistan | 1 | 2017 | 19 | 1931 | 16 | 700 |
| Baluchistan | 2 | 2017 | 19 | 1952 | 15 | 700 |
| Baluchistan | 1 | 2018 | 19 | 1961 | 16 | 867 |
| Baluchistan | 2 | 2018 | 19 | 2040 | 16 | 769 |
| Baluchistan | 1 | 2019 | 19 | 2078 | 15 | 759 |
| Baluchistan | 2 | 2019 | 19 | 2204 | 17 | 790 |
| Baluchistan | 1 | 2020 | 19 | 2084 | 15 | 785 |
| Baluchistan | 2 | 2020 | 19 | 2140 | 16 | 819 |
| Faisalabad | 1 | 2017 | 41 | 5415 | 40 | 2117 |
| Faisalabad | 2 | 2017 | 41 | 5739 | 40 | 2141 |
| Faisalabad | 1 | 2018 | 41 | 5807 | 38 | 2133 |
| Faisalabad | 2 | 2018 | 41 | 5937 | 36 | 2151 |
| Faisalabad | 1 | 2019 | 41 | 6113 | 38 | 2372 |
| Faisalabad | 2 | 2019 | 41 | 6159 | 39 | 2536 |
| Faisalabad | 1 | 2020 | 40 | 5616 | 37 | 2293 |
| Faisalabad | 2 | 2020 | 42 | 6377 | 39 | 2362 |
| Karachi | 1 | 2017 | 137 | 18409 | 120 | 6501 |
| Karachi | 2 | 2017 | 137 | 18668 | 121 | 6489 |
| Karachi | 1 | 2018 | 138 | 19053 | 123 | 6708 |
| Karachi | 2 | 2018 | 138 | 19436 | 124 | 6554 |
| Karachi | 1 | 2019 | 138 | 19956 | 121 | 7331 |
| Karachi | 2 | 2019 | 137 | 19680 | 117 | 6761 |
| Karachi | 1 | 2020 | 133 | 19150 | 112 | 6614 |
| Karachi | 2 | 2020 | 137 | 19410 | 120 | 6771 |
| Lahore | 1 | 2017 | 129 | 17419 | 107 | 4621 |
| Lahore | 2 | 2017 | 129 | 17879 | 105 | 4629 |
| Lahore | 1 | 2018 | 129 | 17931 | 107 | 4703 |
| Lahore | 2 | 2018 | 129 | 18282 | 107 | 4915 |
| Lahore | 1 | 2019 | 129 | 18319 | 109 | 5176 |
| Lahore | 2 | 2019 | 128 | 18408 | 105 | 5047 |
| Lahore | 1 | 2020 | 126 | 17439 | 101 | 4869 |
| Lahore | 2 | 2020 | 127 | 18123 | 101 | 5003 |
| Multan | 1 | 2017 | 47 | 7160 | 40 | 2702 |
| Multan | 2 | 2017 | 47 | 7299 | 39 | 2779 |
| Multan | 1 | 2018 | 46 | 7103 | 38 | 2546 |
| Multan | 2 | 2018 | 46 | 7129 | 38 | 2573 |
| Multan | 1 | 2019 | 46 | 7276 | 39 | 2630 |
| Multan | 2 | 2019 | 47 | 7738 | 43 | 2723 |
| Multan | 1 | 2020 | 47 | 7630 | 38 | 2843 |
| Multan | 2 | 2020 | 47 | 7787 | 40 | 2912 |
| Peshawar | 1 | 2017 | 46 | 5486 | 40 | 1663 |
| Peshawar | 2 | 2017 | 46 | 5891 | 42 | 1761 |
| Peshawar | 1 | 2018 | 46 | 5964 | 41 | 1723 |
| Peshawar | 2 | 2018 | 46 | 6142 | 43 | 1958 |
| Peshawar | 1 | 2019 | 46 | 6256 | 44 | 2022 |
| Peshawar | 2 | 2019 | 46 | 6487 | 42 | 2111 |
| Peshawar | 1 | 2020 | 46 | 6363 | 42 | 2081 |
| Peshawar | 2 | 2020 | 46 | 6335 | 44 | 1965 |
| Rawalpindi | 1 | 2017 | 63 | 8048 | 56 | 2248 |
| Rawalpindi | 2 | 2017 | 63 | 8217 | 55 | 2597 |
| Rawalpindi | 1 | 2018 | 63 | 8312 | 55 | 2719 |
| Rawalpindi | 2 | 2018 | 63 | 8457 | 57 | 2935 |
| Rawalpindi | 1 | 2019 | 63 | 8655 | 56 | 2855 |
| Rawalpindi | 2 | 2019 | 64 | 8953 | 55 | 3026 |
| Rawalpindi | 1 | 2020 | 64 | 9007 | 56 | 2948 |
| Rawalpindi | 2 | 2020 | 64 | 9051 | 55 | 3008 |
| Sindh | 1 | 2017 | 58 | 8799 | 55 | 3485 |
| Sindh | 2 | 2017 | 58 | 8871 | 53 | 3519 |
| Sindh | 1 | 2018 | 58 | 8932 | 53 | 3610 |
| Sindh | 2 | 2018 | 58 | 9018 | 53 | 3556 |
| Sindh | 1 | 2019 | 58 | 9058 | 52 | 3535 |
| Sindh | 2 | 2019 | 58 | 9190 | 54 | 3564 |
| Sindh | 1 | 2020 | 56 | 8930 | 51 | 3605 |
| Sindh | 2 | 2020 | 58 | 9225 | 53 | 3634 |

B. EGYPT
| Region | Quarter/<br>Term | Year | Total number of<br>doctors<br>submitting<br>records of health<br>consultations for<br>any condition | Number of<br>records of<br>health<br>consultations<br>submitted | Number of<br>doctors<br>submitting<br>records of health<br>consultations for<br>common<br>infection-related<br>visits | Number of<br>records of<br>health<br>consultations<br>for common<br>infection-related visits |
| --- | --- | --- | --- | --- | --- | --- |
| Alexandria | 1 | 2017 | 83 | 3111 | 58 | 683 |
| Alexandria | 2 | 2017 | 83 | 3059 | 58 | 654 |
| Alexandria | 3 | 2017 | 86 | 3321 | 61 | 683 |
| Alexandria | 4 | 2017 | 86 | 3248 | 65 | 689 |
| Alexandria | 1 | 2018 | 86 | 1848 | 57 | 378 |
| Alexandria | 2 | 2018 | 86 | 3249 | 62 | 674 |
| Alexandria | 3 | 2018 | 86 | 3267 | 64 | 684 |
| Alexandria | 4 | 2018 | 86 | 3550 | 64 | 743 |
| Alexandria | 1 | 2019 | 86 | 3489 | 65 | 743 |
| Alexandria | 2 | 2019 | 86 | 3496 | 66 | 717 |
| Alexandria | 3 | 2019 | 85 | 3398 | 54 | 704 |
| Alexandria | 4 | 2019 | 85 | 3333 | 58 | 619 |
| Alexandria | 1 | 2020 | 85 | 2923 | 55 | 534 |
| Alexandria | 2 | 2020 | 85 | 2594 | 54 | 472 |
| Alexandria | 3 | 2020 | 85 | 2664 | 49 | 460 |
| Alexandria | 4 | 2020 | 85 | 3056 | 52 | 552 |
| Alexandria | 1 | 2021 | 85 | 3150 | 50 | 512 |
| Alexandria | 2 | 2021 | 85 | 3325 | 53 | 577 |
| Alexandria | 3 | 2021 | 85 | 3245 | 51 | 576 |
| Cairo - Giza | 1 | 2017 | 211 | 9116 | 148 | 1854 |
| Cairo - Giza | 2 | 2017 | 213 | 9277 | 150 | 1839 |
| Cairo - Giza | 3 | 2017 | 222 | 9287 | 154 | 1752 |
| Cairo - Giza | 4 | 2017 | 224 | 9425 | 157 | 1809 |
| Cairo - Giza | 1 | 2018 | 222 | 4818 | 126 | 992 |
| Cairo - Giza | 2 | 2018 | 224 | 9537 | 157 | 1834 |
| Cairo - Giza | 3 | 2018 | 223 | 9363 | 154 | 1697 |
| Cairo - Giza | 4 | 2018 | 224 | 10026 | 157 | 1911 |
| Cairo - Giza | 1 | 2019 | 224 | 10204 | 159 | 1964 |
| Cairo - Giza | 2 | 2019 | 224 | 10330 | 152 | 1977 |
| Cairo - Giza | 3 | 2019 | 253 | 11672 | 171 | 2117 |
| Cairo - Giza | 4 | 2019 | 253 | 10707 | 175 | 1922 |
| Cairo - Giza | 1 | 2020 | 253 | 9672 | 177 | 1776 |
| Cairo - Giza | 2 | 2020 | 251 | 7495 | 159 | 1265 |
| Cairo - Giza | 3 | 2020 | 252 | 8020 | 165 | 1225 |
| Cairo - Giza | 4 | 2020 | 253 | 9650 | 159 | 1553 |
| Cairo - Giza | 1 | 2021 | 253 | 9474 | 157 | 1370 |
| Cairo - Giza | 2 | 2021 | 252 | 9607 | 156 | 1423 |
| Cairo - Giza | 3 | 2021 | 253 | 9365 | 161 | 1300 |
| Delta/Canal Zone | 1 | 2017 | 145 | 5722 | 107 | 1261 |
| Delta/Canal Zone | 2 | 2017 | 145 | 5759 | 109 | 1243 |
| Delta/Canal Zone | 3 | 2017 | 148 | 5980 | 111 | 1243 |
| Delta/Canal Zone | 4 | 2017 | 148 | 6056 | 113 | 1311 |
| Delta/Canal Zone | 1 | 2018 | 147 | 3208 | 89 | 726 |
| Delta/Canal Zone | 2 | 2018 | 148 | 5938 | 99 | 1187 |
| Delta/Canal Zone | 3 | 2018 | 148 | 6139 | 106 | 1220 |
| Delta/Canal Zone | 4 | 2018 | 148 | 6304 | 106 | 1317 |
| Delta/Canal Zone | 1 | 2019 | 148 | 6352 | 103 | 1274 |
| Delta/Canal Zone | 2 | 2019 | 148 | 6398 | 105 | 1331 |
| Delta/Canal Zone | 3 | 2019 | 207 | 8565 | 154 | 1796 |
| Delta/Canal Zone | 4 | 2019 | 206 | 8157 | 151 | 1751 |
| Delta/Canal Zone | 1 | 2020 | 207 | 7653 | 144 | 1541 |
| Delta/Canal Zone | 2 | 2020 | 207 | 6412 | 141 | 1314 |
| Delta/Canal Zone | 3 | 2020 | 207 | 6379 | 143 | 1189 |
| Delta/Canal Zone | 4 | 2020 | 207 | 7786 | 148 | 1457 |
| Delta/Canal Zone | 1 | 2021 | 207 | 8089 | 145 | 1355 |
| Delta/Canal Zone | 2 | 2021 | 207 | 8371 | 147 | 1452 |
| Delta/Canal Zone | 3 | 2021 | 207 | 8575 | 151 | 1522 |
| Upper Egypt | 1 | 2017 | 84 | 3683 | 66 | 949 |
| Upper Egypt | 2 | 2017 | 84 | 3816 | 65 | 996 |
| Upper Egypt | 3 | 2017 | 87 | 4012 | 66 | 953 |
| Upper Egypt | 4 | 2017 | 87 | 3923 | 63 | 920 |
| Upper Egypt | 1 | 2018 | 87 | 2259 | 60 | 547 |
| Upper Egypt | 2 | 2018 | 87 | 3762 | 62 | 910 |
| Upper Egypt | 3 | 2018 | 87 | 3840 | 64 | 846 |
| Upper Egypt | 4 | 2018 | 87 | 4077 | 64 | 977 |
| Upper Egypt | 1 | 2019 | 87 | 4025 | 66 | 938 |
| Upper Egypt | 2 | 2019 | 87 | 4088 | 66 | 969 |
| Upper Egypt | 3 | 2019 | 110 | 5199 | 81 | 1279 |
| Upper Egypt | 4 | 2019 | 110 | 5287 | 81 | 1234 |
| Upper Egypt | 1 | 2020 | 110 | 4583 | 82 | 1009 |
| Upper Egypt | 2 | 2020 | 110 | 4145 | 80 | 870 |
| Upper Egypt | 3 | 2020 | 110 | 4420 | 82 | 899 |
| Upper Egypt | 4 | 2020 | 110 | 4930 | 80 | 1088 |
| Upper Egypt | 1 | 2021 | 110 | 5081 | 78 | 1024 |
| Upper Egypt | 2 | 2021 | 110 | 5294 | 78 | 1083 |
| Upper Egypt | 3 | 2021 | 110 | 5308 | 80 | 1071 |

C. INDONESIA
| Region | Quarter/<br>Term | Year | Total number of<br>doctors<br>submitting<br>records of health<br>consultations for<br>any condition | Number of<br>records of<br>health<br>consultations<br>submitted | Number of doctors<br>submitting records<br>of health<br>consultations for<br>common infection-related visits | Number of<br>records of<br>health<br>consultations<br>for common<br>infection-related visits |
| --- | --- | --- | --- | --- | --- | --- |
| Central Java | 1 | 2017 | 129 | 11461 | 104 | 2244 |
| Central Java | 2 | 2017 | 124 | 11200 | 103 | 2130 |
| Central Java | 1 | 2018 | 122 | 10986 | 103 | 2096 |
| Central Java | 2 | 2018 | 115 | 10348 | 91 | 1999 |
| Central Java | 1 | 2019 | 120 | 11195 | 98 | 2247 |
| East<br>Indonesia | 1 | 2017 | 85 | 8064 | 75 | 1983 |
| East<br>Indonesia | 2 | 2017 | 84 | 8125 | 75 | 1949 |
| East<br>Indonesia | 1 | 2018 | 77 | 7339 | 64 | 1929 |
| East<br>Indonesia | 2 | 2018 | 91 | 8770 | 82 | 2353 |
| East<br>Indonesia | 1 | 2019 | 96 | 9327 | 86 | 2508 |
| East Java | 1 | 2017 | 135 | 11304 | 103 | 2154 |
| East Java | 2 | 2017 | 136 | 12710 | 97 | 2373 |
| East Java | 1 | 2018 | 140 | 13028 | 103 | 2515 |
| East Java | 2 | 2018 | 134 | 12583 | 98 | 2107 |
| East Java | 1 | 2019 | 133 | 12622 | 99 | 2377 |
| Jakarta | 1 | 2017 | 259 | 24223 | 199 | 4964 |
| Jakarta | 2 | 2017 | 266 | 25095 | 211 | 5100 |
| Jakarta | 1 | 2018 | 280 | 26426 | 219 | 5130 |
| Jakarta | 2 | 2018 | 280 | 26704 | 209 | 5183 |
| Jakarta | 1 | 2019 | 276 | 26572 | 210 | 5655 |
| Sumatra | 1 | 2017 | 180 | 17307 | 154 | 3615 |
| Sumatra | 2 | 2017 | 167 | 16167 | 134 | 3381 |
| Sumatra | 1 | 2018 | 167 | 16105 | 137 | 3314 |
| Sumatra | 2 | 2018 | 171 | 16444 | 140 | 3243 |
| Sumatra | 1 | 2019 | 170 | 16475 | 136 | 3263 |
| West Java | 1 | 2017 | 156 | 14487 | 121 | 3379 |
| West Java | 2 | 2017 | 160 | 15010 | 130 | 3471 |
| West Java | 1 | 2018 | 159 | 14832 | 125 | 3531 |
| West Java | 2 | 2018 | 148 | 13839 | 116 | 3366 |
| West Java | 1 | 2019 | 138 | 13007 | 109 | 3248 |

**Appendix 2.** List of ICD-10 code corresponding to common acute infection diagnoses included in the study

| ICD code | Diagnosis | Diagnosis groups |
| --- | --- | --- |
| R50 | Fever of other and unknown origin | Fever |
| A00 | Cholera | Gastrointestinal infections |
| A01 | Typhoid and paratyphoid fevers | Gastrointestinal infections |
| A02 | Other salmonella infections | Gastrointestinal infections |
| A03 | Shigellosis | Gastrointestinal infections |
| A04 | Other bacterial intestinal infections | Gastrointestinal infections |
| A05 | Other bacterial foodborne intoxications, not elsewhere classified | Gastrointestinal infections |
| A06 | Amoebiasis | Gastrointestinal infections |
| A07 | Other protozoal intestinal diseases | Gastrointestinal infections |
| A08 | Viral and other specified intestinal infections | Gastrointestinal infections |
| A09 | Other gastroenteritis and colitis of infectious and unspecified origin | Gastrointestinal infections |
| H60.0 | Abscess of external ear | Mastoiditis and other ear infection complications |
| H60.1 | Cellulitis of external ear | Mastoiditis and other ear infection complications |
| H60.2 | Malignant otitis externa | Mastoiditis and other ear infection complications |
| H60.8 | Other otitis externa | Mastoiditis and other ear infection complications |
| H60.9 | Otitis externa, unspecified | Mastoiditis and other ear infection complications |
| H60.3 | Other infective otitis externa | Mastoiditis and other ear infection complications |
| H62.0 | Otitis externa in bacterial diseases classified elsewhere | Mastoiditis and other ear infection complications |
| H62.3 | Otitis externa in other infectious and parasitic diseases classified elsewhere | Mastoiditis and other ear infection complications |
| H62.4 | Otitis externa in other diseases classified elsewhere | Mastoiditis and other ear infection complications |
| H65.0 | Acute serious otitis media | Mastoiditis and other ear infection complications |
| H65.1 | Other acute nonsuppurative otitis media | Mastoiditis and other ear infection complications |
| H65.9 | Nonsuppurative otitis media, unspecified | Mastoiditis and other ear infection complications |
| H66.0 | Acute suppurative otitis media | Mastoiditis and other ear infection complications |
| H66.4 | Suppurative otitis media, unspecified | Mastoiditis and other ear infection complications |
| H66.9 | Otitis media, unspecified | Mastoiditis and other ear infection complications |
| H67.0 | Otitis media, unspecified | Mastoiditis and other ear infection complications |
| H67.8 | Otitis media in bacterial diseases classified elsewhere | Mastoiditis and other ear infection complications |
| H70 | Mastoiditis and related conditions | Mastoiditis and other ear infection complications |
| H74 | Other disorders of middle ear mastoid | Mastoiditis and other ear infection complications |
| H75 | Other disorders of middle ear and mastoid in diseases classified elsewhere | Mastoiditis and other ear infection complications |
| J12 | Viral pneumonia, not elsewhere classified | Acute lower RTIs |
| J13 | Pneumonia due to *Streptococcus pneumoniae* | Acute lower RTIs |
| J14 | Pneumonia due to *Haemophilus influenzae* | Acute lower RTIs |
| J15 | Bacterial pneumonia, not elsewhere classified | Acute lower RTIs |
| J16 | Pneumonia due to other infectious organisms, not elsewhere classified | Acute lower RTIs |
| J17 | Pneumonia in diseases classified elsewhere | Acute lower RTIs |
| J18 | Pneumonia, organism unspecified | Acute lower RTIs |
| J20 | Acute bronchitis | Acute lower RTIs |
| J21 | Acute bronchiolitis | Acute lower RTIs |
| J22 | Unspecified acute lower respiratory infection | Acute lower RTIs |
| J44.0 | COPD with acute lower respiratory tract infection | Acute lower RTIs |
| J40 | Bronchitis, not specified as acute or chronic | Acute lower RTIs |
| J44.1 | COPD with acute exacerbation, unspecified | Acute lower RTIs |
| A36.0 | Pharyngeal diphtheria | Acute upper RTIs |
| A36.1 | Nasopharyngeal diphtheria | Acute upper RTIs |
| A36.2 | Laryngeal diphtheria | Acute upper RTIs |
| J00 | Acute nasopharyngitis | Acute upper RTIs |
| J01 | Acute sinusitis | Acute upper RTIs |
| J02 | Acute pharyngitis | Acute upper RTIs |
| J03 | Acute tonsillitis | Acute upper RTIs |
| J04 | Acute laryngitis and tracheitis | Acute upper RTIs |
| J05 | Acute obstructive laryngitis [croup] and epiglottitis | Acute upper RTIs |
| J06 | Acute upper respiratory infections of multiple and unspecified sites | Acute upper RTIs |
| J36 | Peritonsillar abscess | Acute upper RTIs |
| J39.0 | Retropharyngeal and parapharyngeal abscess | Acute upper RTIs |
| J39.1 | Other abscess of pharynx | Acute upper RTIs |
| J10 | Influenza due to identified seasonal influenza virus | Influenza |
| J11 | Influenza, virus not identified | Influenza |
| R05 | Cough | Other respiratory acute symptoms |
| R07.0 | Pain in throat | Other respiratory acute symptoms |
| R07.3 | Chest pain on breathing | Other respiratory acute symptoms |
| R07.4 | Chest pain, unspecified | Other respiratory acute symptoms |
| R09.3 | Abnormal sputum | Other respiratory acute symptoms |
| J98.7 | Respiratory infections, not elsewhere classified (Respiratory (tract) infections not specified as acute, chronic, lower, or upper) | Other RTIs |
| J98.8 | Other specified respiratory disorders | Other RTIs |
| A36.3 | Cutaneous diphtheria | Skin infections |
| A46 | Erysipelas | Skin infections |
| K12.2 | Cellulitis and abscess of mouth | Skin infections |
| L00 | Staphylococcal scalded skin syndrome | Skin infections |
| L01 | Impetigo | Skin infections |
| L02 | Cutaneous abscess, furuncle and carbuncle | Skin infections |
| L03 | Cellulitis | Skin infections |
| L04 | Acute lymphadenitis of face, head and neck | Skin infections |
| L08.0 | Pyoderma (excl pyoderma gangrenosum) | Skin infections |
| L08.8 | Other spec local infections of skin and subcutaneous tissue | Skin infections |
| L08.9 | Local infection of skin and subcutaneous tissue unspecified | Skin infections |
| N30.0 | Cystitis | UTI-cystitis |
| N30.9 | Cystitis, unspecified | UTI-cystitis |
| N10 | Acute tubulo-interstitial nephritis | UTI-others |
| N12 | Tubulo-interstitial nephritis, not specified as acute or chronic | UTI-others |
| N13.6 | Pyenephrosis | UTI-others |
| N15.1 | Renal and perinephric abscess | UTI-others |
| N15.9 | Renal tubule-interstitial disease, unspecified | UTI-others |
| N16.0 | Renal tubule-interstitial disorders in infectious and parasitic diseases | UTI-others |
| N30.8 | Other cystitis - abscess of bladder | UTI-others |
| N39.0 | Urinary tract infection, site not specified | UTI-others |
| N41.0 | Acute prostatitis | UTI-others |
| N41.2 | Abscess of prostate | UTI-others |
| N41.3 | Prostatocystitis | UTI-others |
| N41.9 | Inflammatory disease of prostate, unspecified | UTI-others |
| N45 | Orchitis and epidymitis | UTI-others |
| N73.3 | Female acute pelvic peritonitis | UTI-others |
| N73.5 | Female pelvic peritonitis, unspecified | UTI-others |

**Appendix 3.** List of Antimicrobials and Their Corresponding ATC Class, AWaRe Category, and WHO-EML Classification

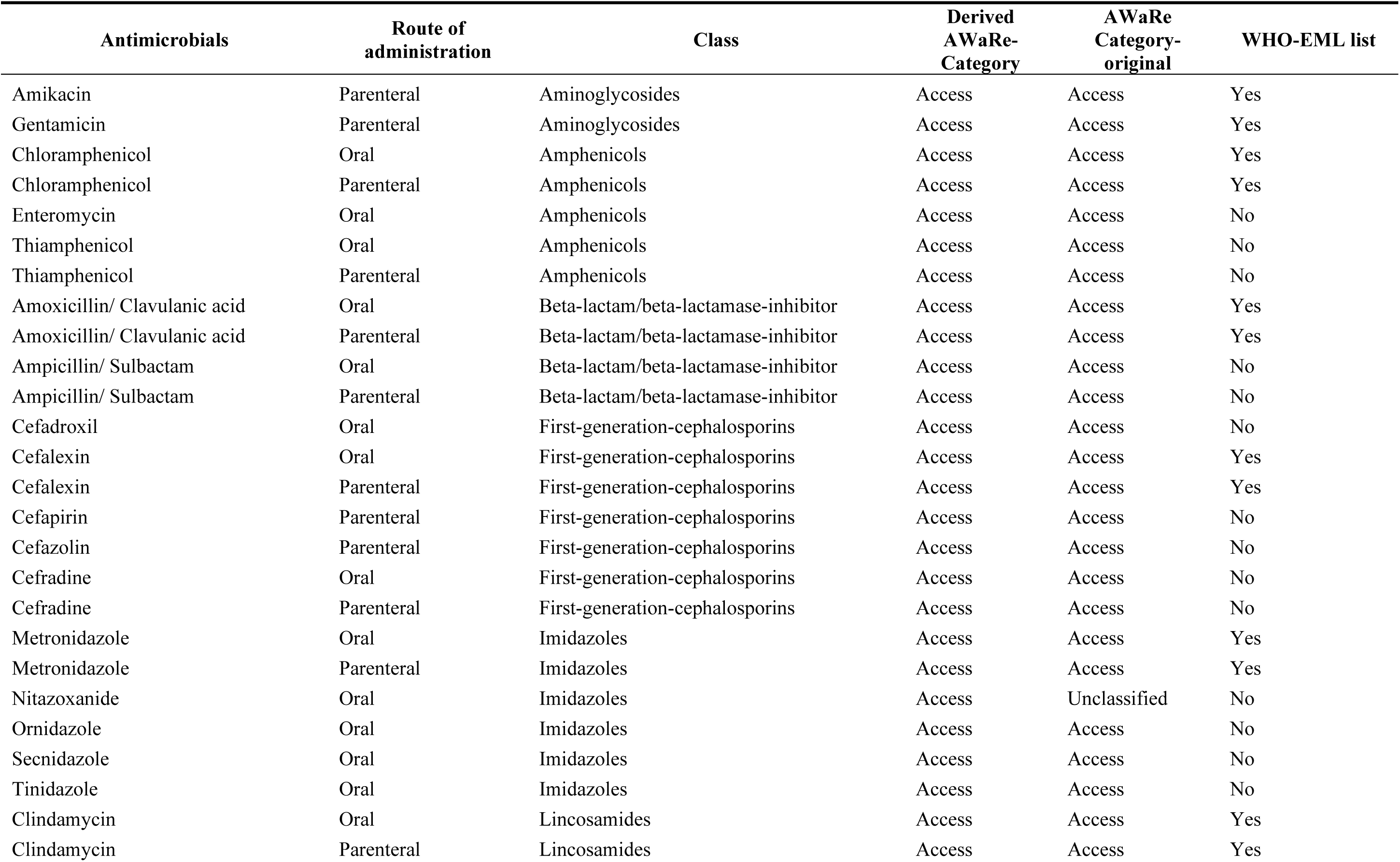
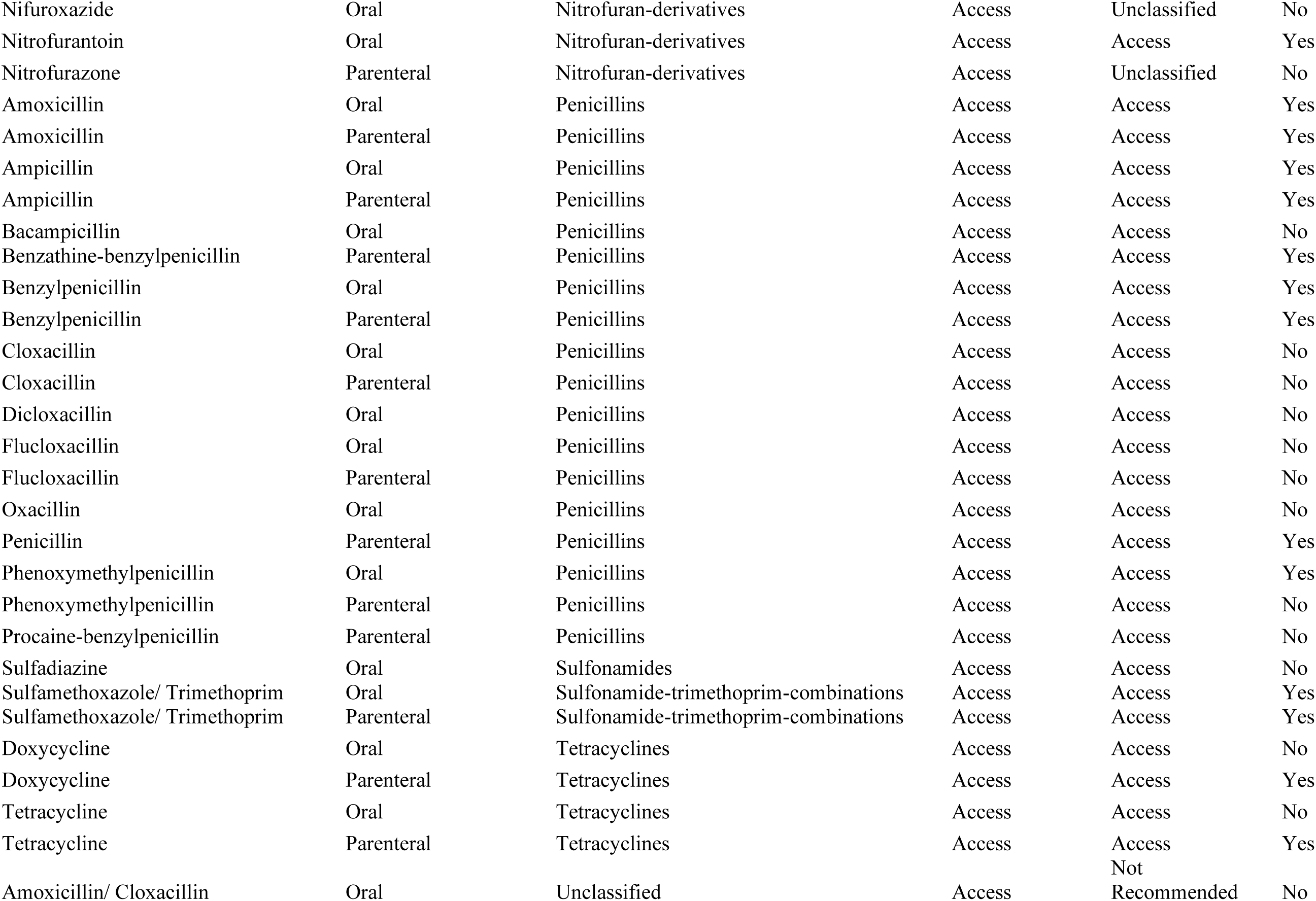
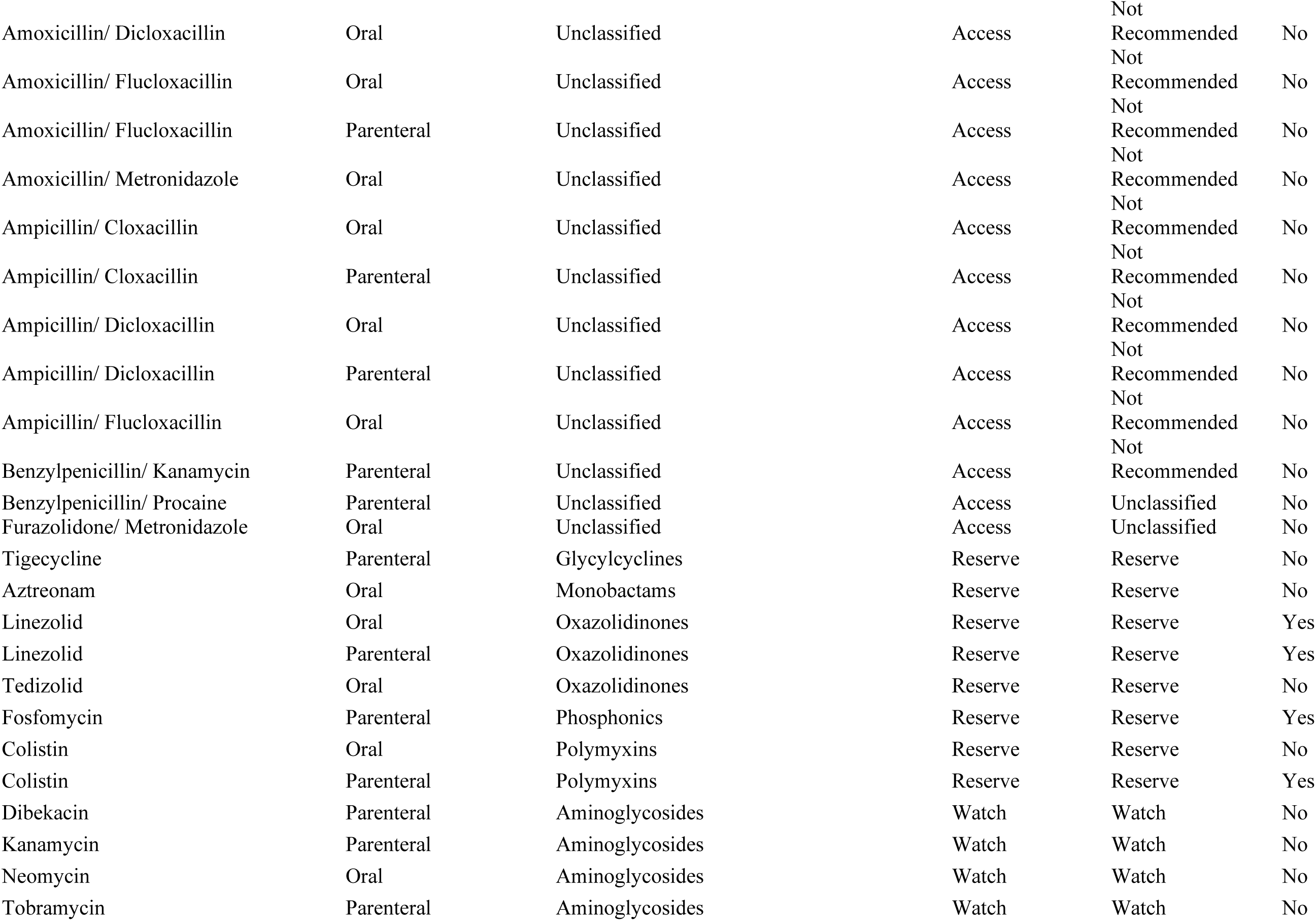
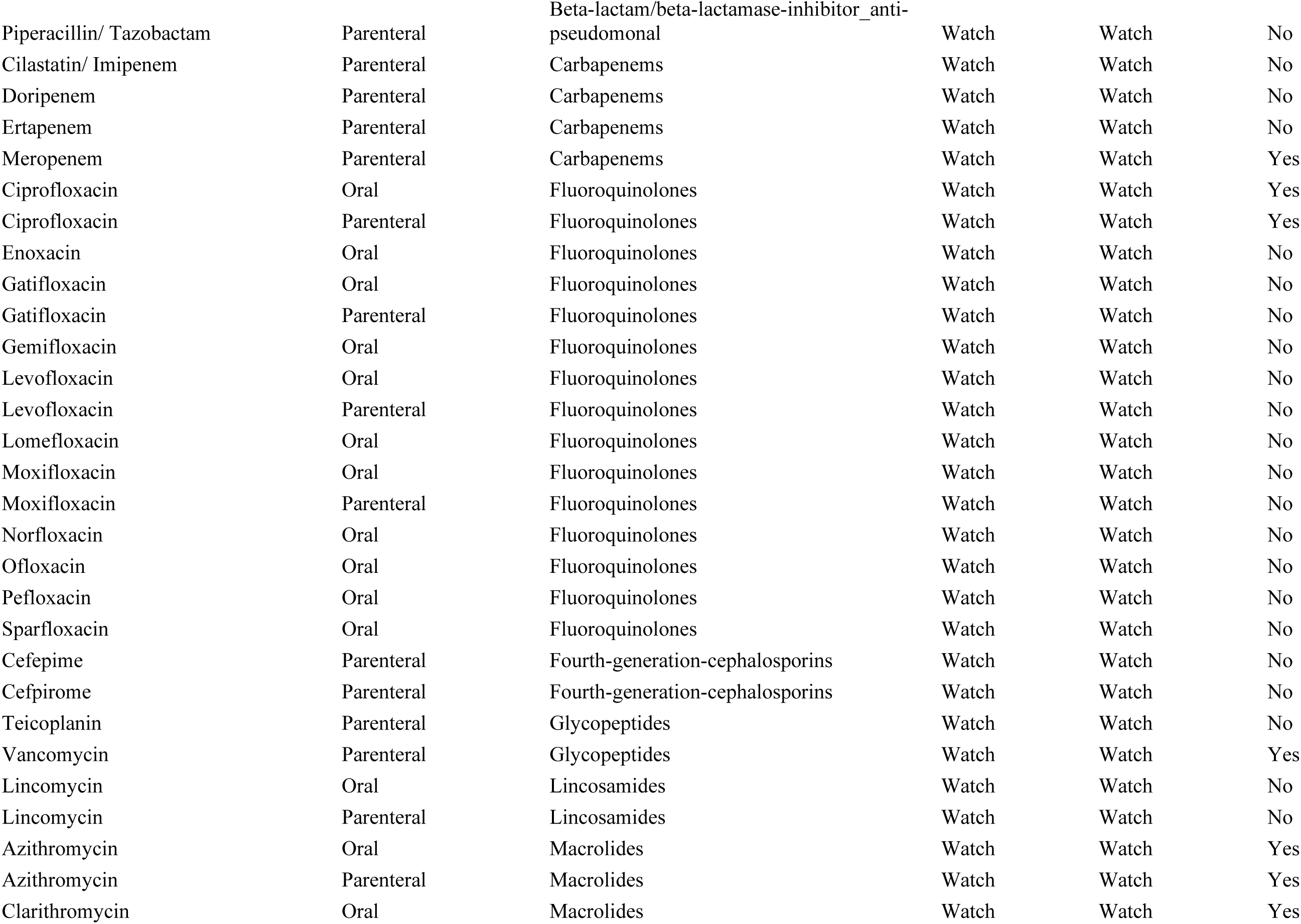
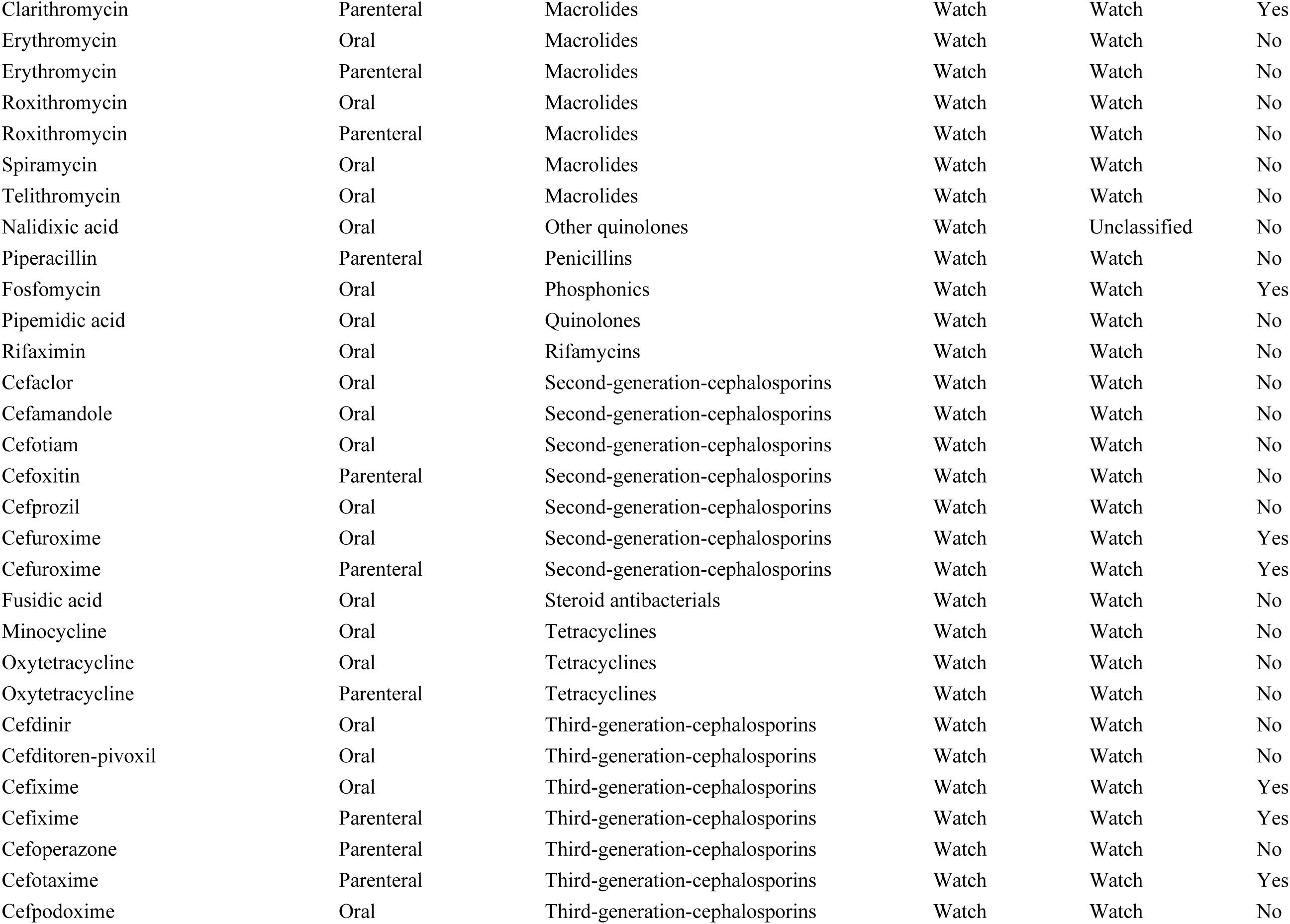
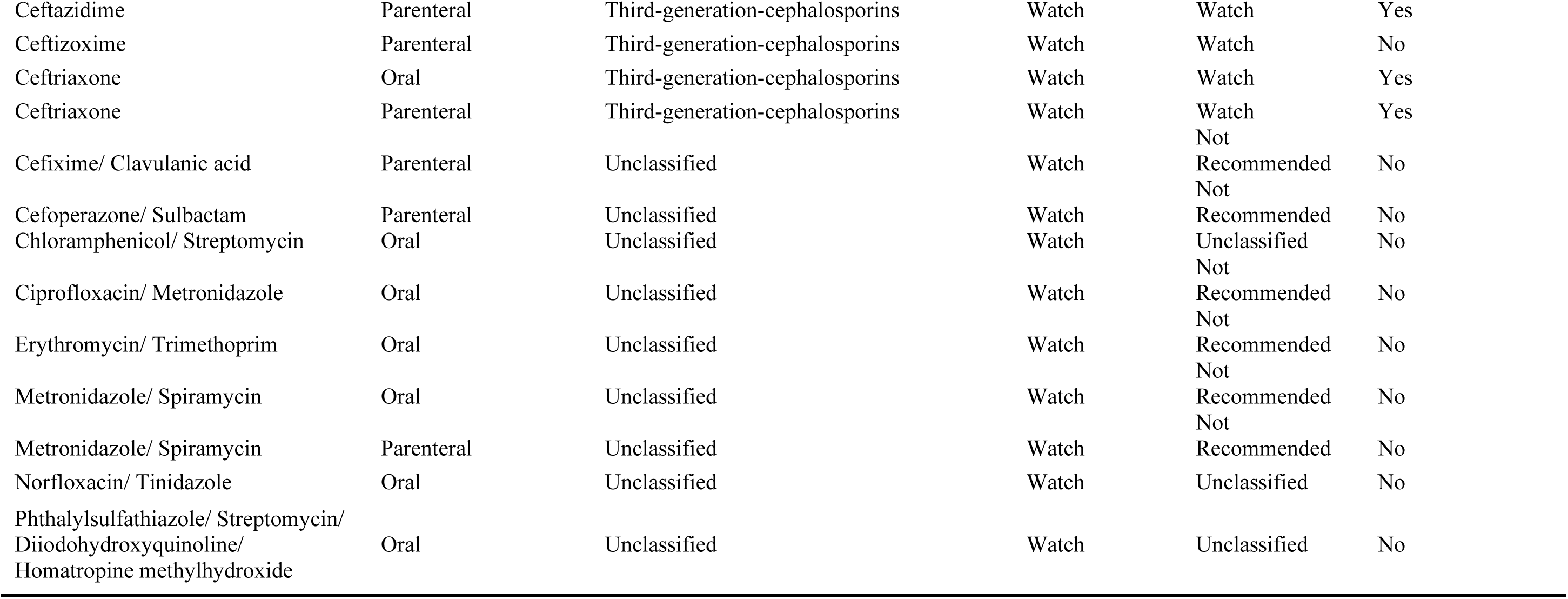

**Appendix 4.** Distribution of antibiotic prescribing by active substances and across countries. Source: Author analysis of IQVIA Medical Data Index (Medical Index of Pakistan (MIP), Egypt Medical Data Index (EMDI), Indonesia Medical Data Index (IMDI)) for the period 2017-2021, reflecting estimates of real world activity. Copyright IQVIA. All rights reserved.

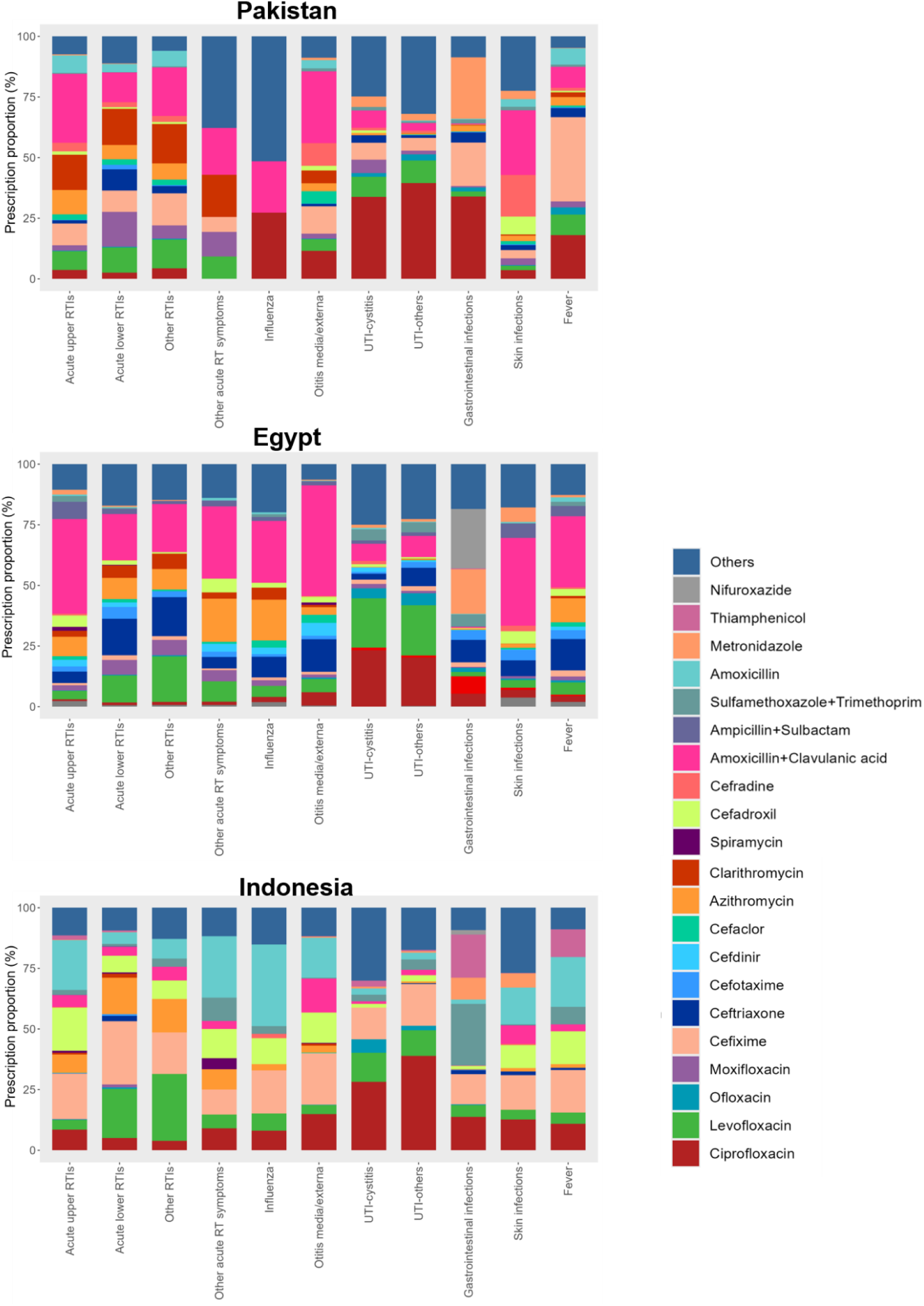

**Appendix 5. WHO AWaRe antibiotic prescription proportional distribution, by prescriber’s speciality and by country.** Source: Author analysis of IQVIA Medical Data Index (Medical Index of Pakistan (MIP), Egypt Medical Data Index (EMDI), Indonesia Medical Data Index (IMDI)) for the period 2017-2021, reflecting estimates of real world activity. Copyright IQVIA. All rights reserved.

|  | Antibiotic prescription proportion | Distributional proportions of WHO-AWaRe antibiotics within the total antibiotic prescription |  |  |
| --- | --- | --- | --- | --- |
|  | N(%) | Accessantibiotics N(%) | Watchantibiotics N(%) | Reserve-antibiotics N(%) |
| **Pakistan** |  |  |  |  |
| General Practitioners | 105321 (84.02) | 31098 (29.53) | 74083 (70.34) | 140 (0.13) |
| Paediatrics | 9808 (75.37) | 3327 (33.92) | 6472 (65.99) | 9 (0.09) |
| Internal Medicine | 8964 (86.96) | 1846 (20.59) | 7084 (79.03) | 34 (0.38) |
| ENT/ Otorhinolaryngology | 6660 (87.36) | 2278 (34.2) | 4364 (65.53) | 18 (0.27) |
| Pulmonology | 5766 (69.76) | 913 (15.83) | 4801 (83.26) | 52 (0.9) |
| Dermatology | 1105 (82.59) | 876 (79.28) | 193 (17.47) | 36 (3.26) |
| Surgery | 2673 (90.61) | 860 (32.17) | 1721 (64.38) | 92 (3.44) |
| Resident Medical Officers | 19841 (79.4) | 6729 (33.91) | 12992 (65.48) | 120 (0.6) |
| Others | 7819 (71.22) | 1546 (19.77) | 6242 (79.83) | 31 (0.4) |
| **Egypt** |  |  |  |  |
| General Practitioners | 12347 (83.34) | 5909 (47.86) | 6383 (51.7) | 55 (0.45) |
| Paediatrics | 14777 (71.74) | 8128 (55) | 6536 (44.23) | 113 (0.76) |
| Internal Medicine | 8575 (79.96) | 3192 (37.22) | 5308 (61.9) | 75 (0.87) |
| ENT/ Otorhinolaryngology | 13217 (87.17) | 7450 (56.37) | 5735 (43.39) | 32 (0.24) |
| Pulmonology | 8887 (84.03) | 2292 (25.79) | 6223 (70.02) | 372 (4.19) |
| Dermatology | 1279 (86.83) | 912 (71.31) | 329 (25.72) | 38 (2.97) |
| Surgery | 2707 (91.08) | 1532 (56.59) | 1064 (39.31) | 111 (4.1) |
| Nephrology/Urology | 4414 (91.39) | 850 (19.26) | 3556 (80.56) | 8 (0.18) |
| Others | 4638 (75.59) | 2275 (49.05) | 2307 (49.74) | 56 (1.21) |
| **Indonesia** |  |  |  |  |
| General Practitioners | 35887 (70.44) | 19110 (53.25) | 16752 (46.68) | 25 (0.07) |
| Paediatrics | 7981 (53.33) | 3997 (50.08) | 3940 (49.37) | 44 (0.55) |
| Internal Medicine | 4419 (77.68) | 1015 (22.97) | 3401 (76.96) | 3 (0.07) |
| ENT/ Otorhinolaryngology | 7444 (73.1) | 3167 (42.54) | 4276 (57.44) | 1 (0.01) |
| Pulmonology | 3017 (74.86) | 460 (15.25) | 2542 (84.26) | 15 (0.5) |
| Dermatology | 1114 (81.85) | 570 (51.17) | 544 (48.83) | 0 (0) |
| Surgery | 1640 (90.71) | 438 (26.71) | 1201 (73.23) | 1 (0.06) |
| Others | 2608 (68.22) | 920 (35.28) | 1682 (64.49) | 6 (0.23) |

**Appendix 6.** Variability in Watch antibiotic prescription proportion among prescribers within the same speciality by country. Source: Author analysis of IQVIA Medical Data Index (Medical Index of Pakistan (MIP), Egypt Medical Data Index (EMDI), Indonesia Medical Data Index (IMDI)) for the period 2017-2021, reflecting estimates of real world activity. Copyright IQVIA. All rights reserved.

PAKISTAN
| Specialisation | Mean (standard deviation) | Coefficient of variation | Median (25<sup>th</sup>-75<sup>th</sup> Percentile) | Interquartile range |
| --- | --- | --- | --- | --- |
| General Practice | 70 (12.4) | 0.18 | 70.3 (63.8-77.6) | 13.8 |
| Paediatrics | 64.3 (18.8) | 0.29 | 63.2 (53.8-74.7) | 20.9 |
| Internal Medicine | 79.1 (10.1) | 0.13 | 79.4 (72.2-83.3) | 11.1 |
| ENT/Otorhinolaryngology | 66 (14.7) | 0.22 | 67.6 (53.6-74.8) | 21.2 |
| Pulmonology | 82.9 (10.7) | 0.13 | 84.1 (74.3-89.8) | 15.5 |
| Dermatology | 23 (17.1) | 0.74 | 21.9 (10-31.8) | 21.9 |
| Surgery | 62.4 (16) | 0.26 | 66.3 (49.1-73.3) | 24.2 |
| Resident Medicine | 68.2 (12.4) | 0.18 | 69.4 (57.7-76) | 18.3 |
| Others | 75.4 (16.7) | 0.22 | 78.8 (67.3-86) | 18.7 |

EGYPT
| Specialisation | Mean (standard deviation) | Coefficient of variation | Median (25<sup>th</sup>-75<sup>th</sup> Percentile) | Interquartile range |
| --- | --- | --- | --- | --- |
| General Practitioners | 51.1 (20.2) | 0.40 | 50 (38-66.3) | 28.2 |
| Paediatrics | 42.3 (17.1) | 0.40 | 40.1 (27.8-53.9) | 26.1 |
| Internal Medicine | 63.6 (16.4) | 0.26 | 66 (52.8-75.5) | 22.7 |
| ENT/Otorhinolaryngology | 44.5 (18.4) | 0.41 | 43.9 (34.2-55) | 20.8 |
| Pulmonology | 74.8 (14.9) | 0.20 | 76.3 (63-87) | 24 |
| Dermatology | 33.3 (23.5) | 0.71 | 28.6 (14.3-43.5) | 29.2 |
| Surgery | 39.5 (19.2) | 0.49 | 40.8 (23.7-55.5) | 31.7 |
| Nephrology/Urology | 79.5 (14.3) | 0.18 | 80.6 (73.5-89.9) | 16.4 |
| Others | 52.8 (23.7) | 0.45 | 52.9 (33.3-73.7) | 40.4 |

INDONESIA
| Specialisation | Mean (standard deviation) | Coefficient of variation | Median (25<sup>th</sup>-75<sup>th</sup> Percentile) | Interquartile range |
| --- | --- | --- | --- | --- |
| General Practitioners | 47.1 (23.3) | 0.50 | 45.8 (29.7-63.2) | 33.6 |
| Paediatrics | 52.7 (25.2) | 0.48 | 55.2 (32.1-72.1) | 39.9 |
| Internal Medicine | 75.6 (18.2) | 0.24 | 80 (65.3-89.4) | 24.1 |
| ENT/Otorhinolaryngology | 56.1 (24.4) | 0.43 | 56.3 (34.2-76.5) | 42.4 |
| Pulmonology | 83 (14.5) | 0.18 | 88.9 (74.6-95) | 20.4 |
| Dermatology | 55.3 (26.6) | 0.48 | 60 (35.4-75) | 39.6 |
| Surgery | 68.9 (21) | 0.31 | 73.9 (57-85.5) | 28.5 |
| Others | 64.5 (26.9) | 0.42 | 73 (41.3-87.5) | 46.2 |

**Appendix 7.** The number and proportion of infectious diagnoses, along with their corresponding categorization indicating the need for antibiotic treatment to manage the disease. Source: Author analysis of IQVIA Medical Data Index (Medical Index of Pakistan (MIP), Egypt Medical Data Index (EMDI), Indonesia Medical Data Index (IMDI)) for the period 2017-2021, reflecting estimates of real world activity. Copyright IQVIA. All rights reserved.

A: always requiring antibiotic; S: sometimes requiring antibiotics; N: never requiring antibiotics.

Source: Chua K-P, Fischer MA, Linder JA. Appropriateness of outpatient antibiotic prescribing among privately insured US patients: ICD-10-CM based cross sectional study. *BMJ* 2019; **364**: k5092.

PAKISTAN
| Infection group | Diagnosis | category | N (%) |
| --- | --- | --- | --- |
| Acute upper RTIs | Acute pharyngitis | S | 17 237 (8.41) |
| Acute upper RTIs | Acute upper respiratory infections of multiple and unspecified sites | N | 15 022 (7.33) |
| Acute upper RTIs | Acute tonsillitis | S | 14 817 (7.23) |
| Acute upper RTIs | Acute sinusitis | S | 3 531 (1.72) |
| Acute upper RTIs | Acute nasopharyngitis | N | 2 565 (1.25) |
| Acute upper RTIs | Acute laryngitis and tracheitis | S | 1 230 (0.6) |
| Acute upper RTIs | Peritonsillar abscess | A | 89 (0.04) |
| Acute upper RTIs | Acute obstructive laryngitis [croup] and epiglottitis | S | 32 (0.02) |
| Acute lower RTIs | Acute bronchitis | N | 6 268 (3.06) |
| Acute lower RTIs | Pneumonia, organism unspecified | A | 4 255 (2.08) |
| Acute lower RTIs | Unspecified acute lower respiratory infection | S | 2 204 (1.08) |
| Acute lower RTIs | COPD with acute exacerbation, unspecified | S | 685 (0.33) |
| Acute lower RTIs | Pneumonia due to *Streptococcus pneumoniae* | A | 237 (0.12) |
| Acute lower RTIs | Acute bronchiolitis | N | 181 (0.09) |
| Acute lower RTIs | Bronchitis, not specified as acute or chronic | N | 139 (0.07) |
| Acute lower RTIs | Bacterial pneumonia, not elsewhere classified | A | 33 (0.02) |
| Influenza | Influenza, virus not identified | S | 13 691 (6.68) |
| Other RTIs | Other specified respiratory disorders | N | 28 600 (13.96) |
| Other respiratory acute symptoms | Cough | N | 43 389 (21.18) |
| Other respiratory acute symptoms | Chest pain, unspecified | N | 2 834 (1.38) |
| Other respiratory acute symptoms | Pain in throat | N | 832 (0.41) |
| Other respiratory acute symptoms | Abnormal sputum | N | 68 (0.03) |
| Other respiratory acute symptoms | Chest pain on breathing | N | 12 (0.01) |
| Mastoiditis and other ear infection complications | Otitis media, unspecified | S | 3 130 (1.53) |
| Mastoiditis and other ear infection complications | Acute suppurative otitis media | S | 1 739 (0.85) |
| Mastoiditis and other ear infection complications | Other infective otitis externa | S | 1 050 (0.51) |
| Mastoiditis and other ear infection complications | Abscess of external ear | S | 573 (0.28) |
| Mastoiditis and other ear infection complications | Nonsuppurative otitis media, unspecified | N | 115 (0.06) |
| Mastoiditis and other ear infection complications | Mastoiditis and related conditions | S | 103 (0.05) |
| Gastrointestinal infections | Other gastroenteritis and colitis of infectious and unspecified origin | S | 32 545 (15.88) |
| Gastrointestinal infections | Typhoid and paratyphoid fevers | A | 12 035 (5.87) |
| Gastrointestinal infections | Amoebiasis | A | 918 (0.45) |
| Gastrointestinal infections | Shigellosis | S | 183 (0.09) |
| Gastrointestinal infections | Other protozoal intestinal diseases | S | 113 (0.06) |
| Gastrointestinal infections | Other bacterial intestinal infections | S | 44 (0.02) |
| Gastrointestinal infections | Cholera | S | 28 (0.01) |
| Gastrointestinal infections | Viral and other specified intestinal infections | S | 11 (0.01) |
| UTI-cystitis | Cystitis, unspecified | S | 766 (0.37) |
| UTI-cystitis | Cystitis | A | 222 (0.11) |
| UTI-others | Urinary tract infection, site not specified | A | 20 572 (10.04) |
| UTI-others | Orchitis and epidymitis | S | 457 (0.22) |
| UTI-others | Acute tubulo-interstitial nephritis | A | 129 (0.06) |
| UTI-others | Acute salpingitis and oophoritis | A | 41 (0.02) |
| UTI-others | Inflammatory disease of prostate, unspecified | S | 25 (0.01) |
| UTI-others | Other cystitis - abscess of bladder | S | 7 (0) |
| Skin infections | Cutaneous abscess, furuncle and carbuncle | S | 3 077 (1.5) |
| Skin infections | Cellulitis | S | 1 619 (0.79) |
| Skin infections | Impetigo | S | 851 (0.42) |
| Skin infections | Local infection of skin and subcutaneous tissue unspecified | S | 770 (0.38) |
| Skin infections | Other spec local infections of skin and subcutaneous tissue | S | 374 (0.18) |
| Skin infections | Acute lymphadenitis | S | 208 (0.1) |
| Skin infections | Pyoderma (excl pyoderma gangrenosum) | S | 103 (0.05) |
| Skin infections | Erysipelas | A | 34 (0.02) |
| Skin infections | Cellulitis and abscess of mouth | A | 11 (0.01) |
| Fever | Fever of other and unknown origin | N | 59 936 (29.25) |
|  | Others |  | 14 (0.01) |

EGYPT
| Infection group | Diagnosis | category | N (%) |
| --- | --- | --- | --- |
| Acute upper RTIs | Acute tonsillitis | S | 9 284 (10.64) |
| Acute upper RTIs | Acute pharyngitis | S | 6 244 (7.15) |
| Acute upper RTIs | Acute nasopharyngitis | N | 5 369 (6.15) |
| Acute upper RTIs | Acute upper respiratory infections of multiple and unspecified sites | N | 2 826 (3.24) |
| Acute upper RTIs | Acute sinusitis | S | 2 356 (2.7) |
| Acute upper RTIs | Acute laryngitis and tracheitis | S | 995 (1.14) |
| Acute upper RTIs | Peritonsillar abscess | A | 143 (0.16) |
| Acute upper RTIs | Acute obstructive laryngitis [croup] and epiglottitis | S | 131 (0.15) |
| Acute lower RTIs | Acute bronchitis | N | 10 099 (11.57) |
| Acute lower RTIs | Bronchitis, not specified as acute or chronic | N | 2 925 (3.35) |
| Acute lower RTIs | Pneumonia, organism unspecified | A | 1 926 (2.21) |
| Acute lower RTIs | Acute bronchiolitis | N | 1 039 (1.19) |
| Acute lower RTIs | COPD with acute exacerbation, unspecified | S | 669 (0.77) |
| Acute lower RTIs | Unspecified acute lower respiratory infection | S | 134 (0.15) |
| Acute lower RTIs | Viral pneumonia, not elsewhere classified | N | 93 (0.11) |
| Acute lower RTIs | COPD with acute lower respiratory tract infection | S | 38 (0.04) |
| Acute lower RTIs | Bacterial pneumonia, not elsewhere classified | A | 17 (0.02) |
| Influenza | Influenza, virus not identified | S | 1 463 (1.68) |
| Other RTIs | Other specified respiratory disorders | N | 1 357 (1.55) |
| Other RTIs | Respiratory infections, not elsewhere classified (Respiratory (tract) infections not specified as acute, chronic, lower, or upper) | S | 13 (0.01) |
| Other respiratory acute symptoms | Cough | N | 4 010 (4.59) |
| Other respiratory acute symptoms | Chest pain, unspecified | N | 1 097 (1.26) |
| Other respiratory acute symptoms | Chest pain on breathing | N | 205 (0.23) |
| Other respiratory acute symptoms | Abnormal sputum | N | 129 (0.15) |
| Mastoiditis and other ear infection complications | Otitis media, unspecified | S | 4 611 (5.28) |
| Mastoiditis and other ear infection complications | Otitis externa, unspecified | S | 1 382 (1.58) |
| Mastoiditis and other ear infection complications | Nonsuppurative otitis media, unspecified | N | 608 (0.7) |
| Mastoiditis and other ear infection complications | Acute suppurative otitis media | S | 175 (0.2) |
| Mastoiditis and other ear infection complications | Suppurative otitis media, unspecified | S | 153 (0.18) |
| Mastoiditis and other ear infection complications | Abscess of external ear | S | 137 (0.16) |
| Mastoiditis and other ear infection complications | Other acute nonsuppurative otitis media | N | 92 (0.11) |
| Mastoiditis and other ear infection complications | Malignant otitis externa | A | 74 (0.08) |
| Mastoiditis and other ear infection complications | Acute serous otitis media | N | 63 (0.07) |
| Mastoiditis and other ear infection complications | Other infective otitis externa | S | 41 (0.05) |
| Mastoiditis and other ear infection complications | Mastoiditis and related conditions | S | 24 (0.03) |
| Mastoiditis and other ear infection complications | Other otitis externa | S | 23 (0.03) |
| Mastoiditis and other ear infection complications | Other disorders of middle ear mastoid | N | 14 (0.02) |
| Gastrointestinal infections | Other gastroenteritis and colitis of infectious and unspecified origin | S | 11 426 (13.09) |
| Gastrointestinal infections | Amoebiasis | A | 835 (0.96) |
| Gastrointestinal infections | Typhoid and paratyphoid fevers | A | 814 (0.93) |
| Gastrointestinal infections | Other protozoal intestinal diseases | S | 181 (0.21) |
| Gastrointestinal infections | Other bacterial intestinal infections | S | 70 (0.08) |
| Gastrointestinal infections | Viral and other specified intestinal infections | S | 46 (0.05) |
| Gastrointestinal infections | Shigellosis | S | 43 (0.05) |
| Gastrointestinal infections | Other bacterial foodborne intoxications, not elsewhere classified | S | 10 (0.01) |
| UTI-cystitis | Cystitis, unspecified | S | 1 513 (1.73) |
| UTI-cystitis | Cystitis | A | 796 (0.91) |
| UTI-others | Urinary tract infection, site not specified | A | 4 831 (5.53) |
| UTI-others | Orchitis and epidymitis | S | 972 (1.11) |
| UTI-others | Inflammatory disease of prostate, unspecified | S | 511 (0.59) |
| UTI-others | Tubulo-interstitial nephritis, not specified as acute or chronic | N | 262 (0.3) |
| UTI-others | Acute prostatitis | A | 227 (0.26) |
| UTI-others | Acute tubulo-interstitial nephritis | A | 117 (0.13) |
| UTI-others | Prostatocystitis | S | 50 (0.06) |
| UTI-others | Salpingitis and oophoritis, unspecified | A | 33 (0.04) |
| UTI-others | Renal tubule-interstitial disease, unspecified | N | 20 (0.02) |
| UTI-others | Other cystitis - abscess of bladder | S | 15 (0.02) |
| UTI-others | Renal and perinephric abscess | A | 14 (0.02) |
| UTI-others | Acute salpingitis and oophoritis | A | 6 (0.01) |
| UTI-others | Pyenephrosis | N | 6 (0.01) |
| Skin infections | Cutaneous abscess, furuncle and carbuncle | S | 2 318 (2.66) |
| Skin infections | Cellulitis | S | 1 652 (1.89) |
| Skin infections | Impetigo | S | 602 (0.69) |
| Skin infections | Local infection of skin and subcutaneous tissue unspecified | S | 318 (0.36) |
| Skin infections | Erysipelas | A | 289 (0.33) |
| Skin infections | Acute lymphadenitis | S | 130 (0.15) |
| Skin infections | Pyoderma (excl pyoderma gangrenosum) | S | 99 (0.11) |
| Skin infections | Staphylococcal scalded skin syndrome | A | 86 (0.1) |
| Skin infections | Cellulitis and abscess of mouth | A | 69 (0.08) |
| Skin infections | Other spec local infections of skin and subcutaneous tissue | S | 39 (0.04) |
| Fever | Fever of other and unknown origin | N | 7 009 (8.03) |
|  | Other |  | 22 (0.03) |

INDONESIA
| Infection group | Diagnosis | Category | N (%) |
| --- | --- | --- | --- |
| Acute upper RTIs | Acute upper respiratory infections of multiple and unspecified sites | N | 26 320 (28.36) |
| Acute upper RTIs | Acute nasopharyngitis | N | 10 468 (11.28) |
| Acute upper RTIs | Acute pharyngitis | S | 8 075 (8.7) |
| Acute upper RTIs | Acute tonsillitis | S | 1 607 (1.73) |
| Acute upper RTIs | Acute laryngitis and tracheitis | S | 441 (0.48) |
| Acute upper RTIs | Peritonsillar abscess | A | 118 (0.13) |
| Acute upper RTIs | Acute sinusitis | S | 112 (0.12) |
| Acute upper RTIs | Acute obstructive laryngitis [croup] and epiglottitis | S | 25 (0.03) |
| Acute upper RTIs | Retropharyngeal and parapharyngeal abscess | A | 10 (0.01) |
| Acute lower RTIs | Pneumonia, organism unspecified | A | 3 118 (3.36) |
| Acute lower RTIs | Bronchitis, not specified as acute or chronic | N | 2 355 (2.54) |
| Acute lower RTIs | Acute bronchitis | N | 826 (0.89) |
| Acute lower RTIs | COPD with acute exacerbation, unspecified | S | 216 (0.23) |
| Acute lower RTIs | Unspecified acute lower respiratory infection | S | 132 (0.14) |
| Acute lower RTIs | Acute bronchiolitis | N | 103 (0.11) |
| Acute lower RTIs | Bacterial pneumonia, not elsewhere classified | A | 27 (0.03) |
| Influenza | Influenza, virus not identified | S | 1 063 (1.15) |
| Other RTIs | Other specified respiratory disorders | N | 464 (0.5) |
| Other respiratory acute symptoms | Cough | N | 1 026 (1.11) |
| Other respiratory acute symptoms | Chest pain, unspecified | N | 325 (0.35) |
| Other respiratory acute symptoms | Abnormal sputum | N | 12 (0.01) |
| Mastoiditis and other ear infection complications | Otitis media, unspecified | S | 2 698 (2.91) |
| Mastoiditis and other ear infection complications | Otitis externa, unspecified | S | 2 617 (2.82) |
| Mastoiditis and other ear infection complications | Nonsuppurative otitis media, unspecified | N | 538 (0.58) |
| Mastoiditis and other ear infection complications | Other infective otitis externa | S | 342 (0.37) |
| Mastoiditis and other ear infection complications | Suppurative otitis media, unspecified | S | 138 (0.15) |
| Mastoiditis and other ear infection complications | Acute suppurative otitis media | S | 119 (0.13) |
| Mastoiditis and other ear infection complications | Abscess of external ear | S | 80 (0.09) |
| Mastoiditis and other ear infection complications | Mastoiditis and related conditions | S | 49 (0.05) |
| Mastoiditis and other ear infection complications | Other acute nonsuppurative otitis media | N | 45 (0.05) |
| Mastoiditis and other ear infection complications | Other otitis externa | S | 12 (0.01) |
| Gastrointestinal infections | Other gastroenteritis and colitis of infectious and unspecified origin | S | 11 730 (12.64) |
| Gastrointestinal infections | Typhoid and paratyphoid fevers | A | 3 486 (3.76) |
| Gastrointestinal infections | Amoebiasis | A | 246 (0.27) |
| Gastrointestinal infections | Shigellosis | S | 62 (0.07) |
| Gastrointestinal infections | Viral and other specified intestinal infections | S | 19 (0.02) |
| Gastrointestinal infections | Other bacterial foodborne intoxications, not elsewhere classified | S | 17 (0.02) |
| Gastrointestinal infections | Other bacterial intestinal infections | S | 17 (0.02) |
| Gastrointestinal infections | Cholera | S | 7 (0.01) |
| UTI-cystitis | Cystitis, unspecified | S | 643 (0.69) |
| UTI-cystitis | Cystitis | A | 87 (0.09) |
| UTI-others | Urinary tract infection, site not specified | A | 4 853 (5.23) |
| UTI-others | Orchitis and epidymitis | S | 215 (0.23) |
| UTI-others | Salpingitis and oophoritis, unspecified | A | 176 (0.19) |
| UTI-others | Tubulo-interstitial nephritis, not specified as acute or chronic | N | 30 (0.03) |
| UTI-others | Acute tubulo-interstitial nephritis | A | 18 (0.02) |
| UTI-others | Inflammatory disease of prostate, unspecified | S | 15 (0.02) |
| Skin infections | Cutaneous abscess, furuncle and carbuncle | S | 2 596 (2.8) |
| Skin infections | Cellulitis | S | 1 291 (1.39) |
| Skin infections | Pyoderma (excl pyoderma gangrenosum) | S | 792 (0.85) |
| Skin infections | Impetigo | S | 666 (0.72) |
| Skin infections | Staphylococcal scalded skin syndrome | A | 280 (0.3) |
| Skin infections | Local infection of skin and subcutaneous tissue unspecified | S | 74 (0.08) |
| Skin infections | Acute lymphadenitis | S | 72 (0.08) |
| Skin infections | Erysipelas | A | 48 (0.05) |
| Skin infections | Cellulitis and abscess of mouth | A | 39 (0.04) |
| Fever | Fever of other and unknown origin | N | 3 966 (4.27) |
|  | Other |  | 39 (0.04) |

**Appendix 8.** The number of antimicrobial agents, antimicrobial products, and suppliers identified in the prescriptions submitted by participating doctors, categorised by WHO-AWaRe classification from each country included in the study. Source: Author analysis of IQVIA Medical Data Index (Medical Index of Pakistan (MIP), Egypt Medical Data Index (EMDI), Indonesia Medical Data Index (IMDI)) for the period 2017-2021, reflecting estimates of real world activity. Copyright IQVIA. All rights reserved.

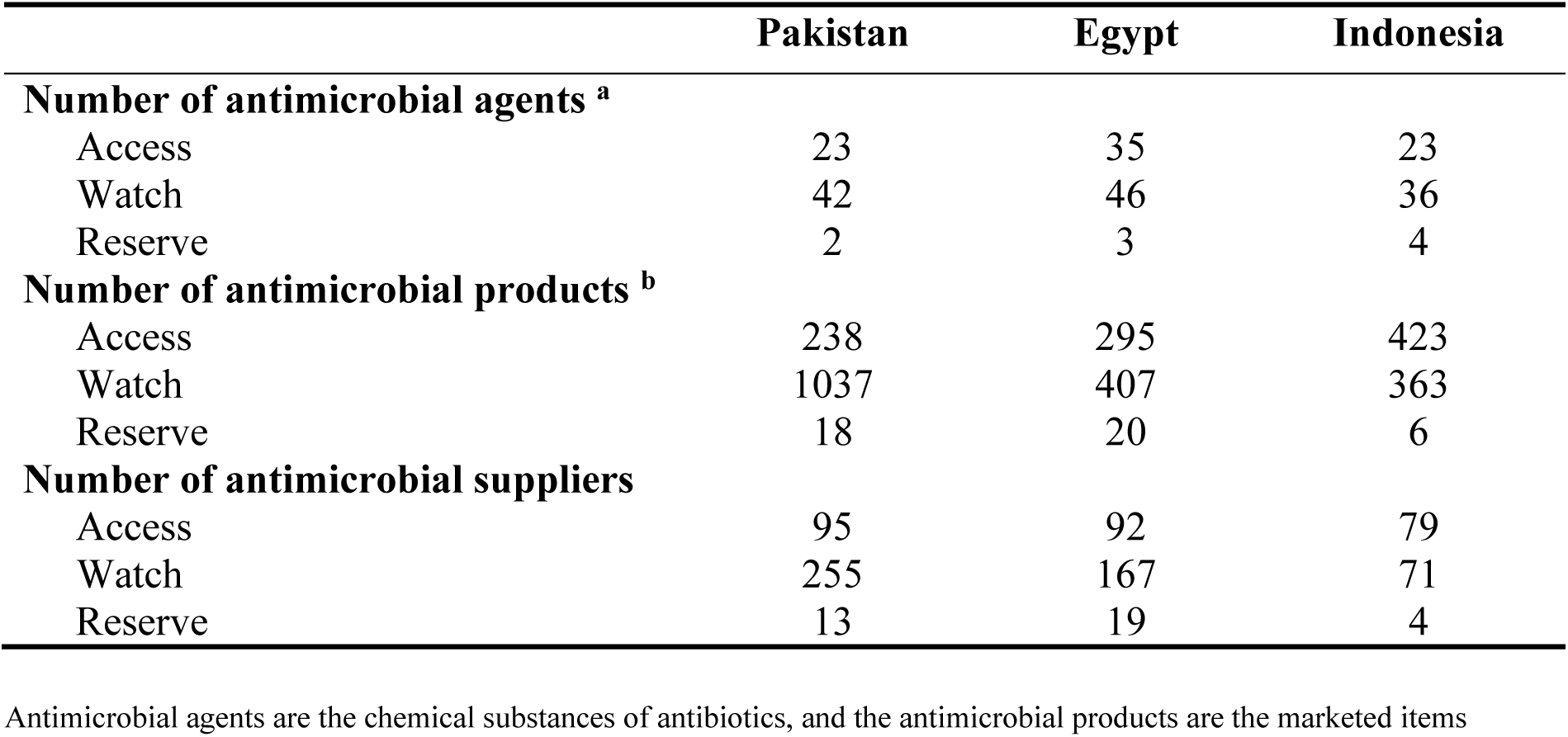

|  | Pakistan | Egypt | Indonesia |
| --- | --- | --- | --- |
| **Number of antimicrobial agents <sup>a</sup>** |  |  |  |
| Access | 23 | 35 | 23 |
| Watch | 42 | 46 | 36 |
| Reserve | 2 | 3 | 4 |
| **Number of antimicrobial products <sup>b</sup>** |  |  |  |
| Access | 238 | 295 | 423 |
| Watch | 1037 | 407 | 363 |
| Reserve | 18 | 20 | 6 |
| **Number of antimicrobial suppliers** |  |  |  |
| Access | 95 | 92 | 79 |
| Watch | 255 | 167 | 71 |
| Reserve | 13 | 19 | 4 |

Antimicrobial agents are the chemical substances of antibiotics, and the antimicrobial products are the marketed items

**Appendix 9.** Factors associated with antibiotic prescribing decisions for patients in Indonesia, with settings classified into doctor’s clinic, governmental hospitals and private hospitals. Source: Author analysis of IQVIA Medical Data Index (Indonesia Medical Data Index (IMDI))), reflecting estimates of real world activity. Copyright IQVIA. All rights reserved.

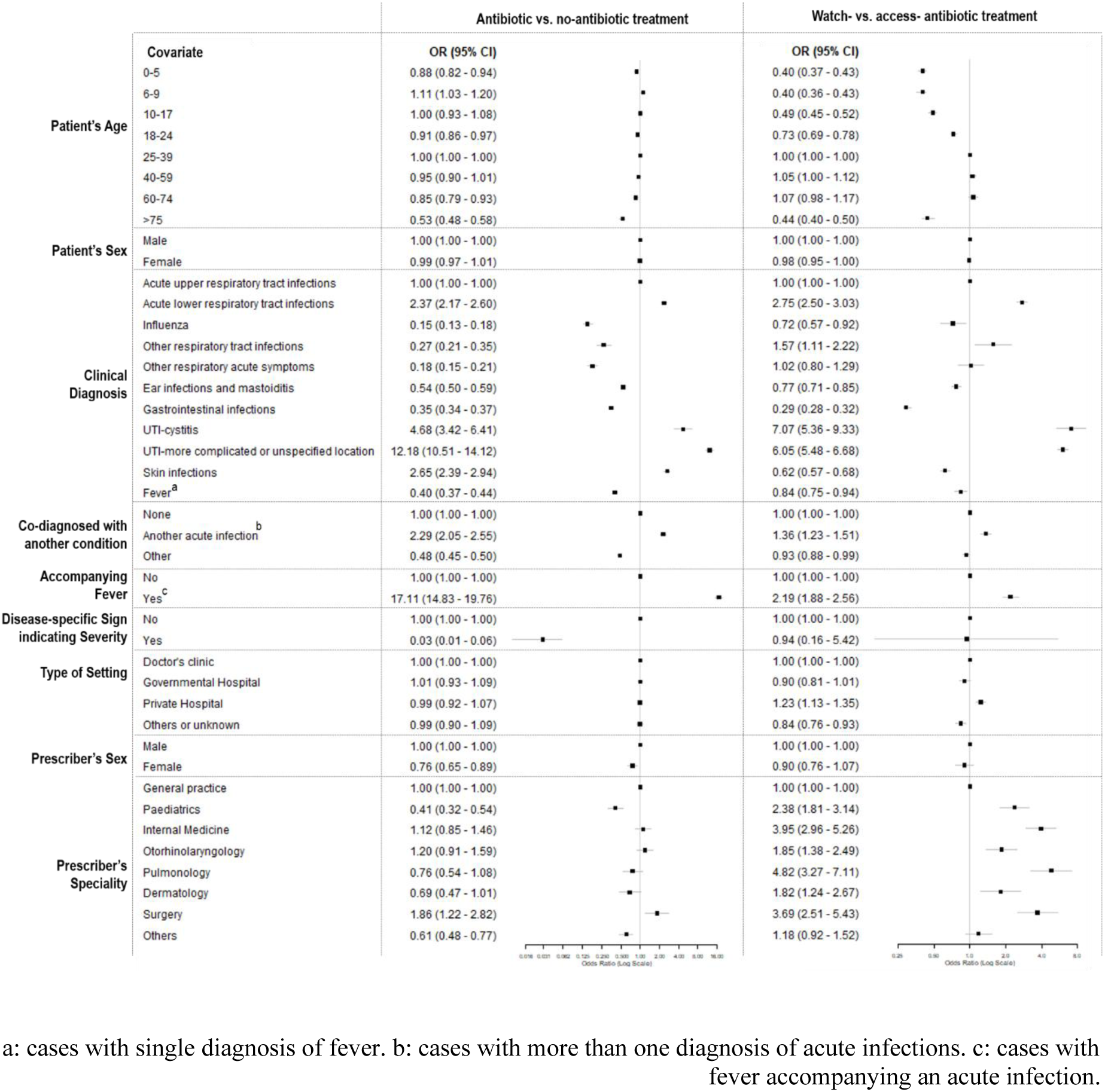

